# Growing up unloved: the enduring consequence of childhood emotional neglect on the qualia of memory and imagination

**DOI:** 10.1101/2022.04.16.22273926

**Authors:** Sinéad L. Mullally, Désirée Grafton-Clarke, Eleanor R Mawson, Matthew Unwin, Molly Stapleton, Kat Webber, Alyson L Dodd, Gillian V Pepper, Derya Cokal, Lucy J Robinson, Peter Gallagher, Stuart Watson

## Abstract

Childhood Adversity (CA) is one of the strongest factors associated with the onset of Major Depressive Disorder (MDD), and both CA and MDD have been linked to altered hippocampal structure/function. The current study aimed to explore the relationships between retrospectively reported childhood emotional neglect (CEN), current wellbeing and depressive symptoms, and a range of hippocampal-dependent cognitive functions i.e., anterograde learning and memory, episodic memory recollection, and imagination (episodic future thinking and scene construction). In two-wave recruitment periods at undergraduate intake 2014-15 (Cohort 1) and 2016-17 (Cohort 2), a combined cohort of n=1485 university students completed online surveys, with n=64 further participating in experimental testing session. As anticipated, higher CEN ratings consistently correlated with poorer current wellbeing and higher depressive symptoms. However, whilst the anticipated relationships between CEN, current wellbeing, and subjectively reported estimates of hippocampal-dependent cognitions were observed in the data reported in the online survey, an unexpectedly circumscribed pattern was observed on formal in-person examination of these cognitive functions. More specifically, higher CEN related to less vivid and less detailed imagined future/scene constructions and with an attenuated sense of presence and emotional valence during these simulations. A similar pattern was not evidence when participants simulated experienced past events (i.e. episodic memories). Current depression scores did not consistently correlate with vividness, detail, or emotional valence. In addition, and contrary to expectation, no relationship between CEN, depressive symptoms, and the spatial coherence of imagined or recollected events was seen. Moreover, neither CEN nor depressive symptoms correlated with many key measures of anterograde memory. Hence, we observed a highly specific constellation of impairment related to CEN when explored on a simulation per simulation basis, that was not obviously linked to altered hippocampal function, indicating that the relationship between CEN, hippocampal function, and subsequent psychopathology may not readily explained by either spatial or mnemonic hippocampal-related deficits. We consider whether the observed experiential differences in the qualia of imagined simulations may represent an important therapeutic target to decrease a CEN-driven latent vulnerability to MDD.

---

***Key words***: childhood adversity, depression, hippocampus, episodic memory, imagination

Childhood adversity (CA) is defined as exposure to abuse or neglect within vital periods of development. CA has been repeatedly demonstrated to be a major risk factor for the development of a variety of mental health problems in adolescence and adulthood, including bipolar disorder (Watson et al., 2014), schizophrenia (Varese et al., 2012), post-traumatic stress disorder (PTSD) (Koenen et al., 2007), anxiety (Young et al., 1997), poor psychological functioning (Raposa et al., 2014; van Vugt et al., 2014), impulse control and substance abuse disorders (Putnam et al., 2013) and, particularly, major depressive disorder (MDD) (Bernet & Stein, 1999; Gibb et al., 2007; Nanni et al., 2012; Phillips et al., 2005; Selous et al., 2019). CA is one of the strongest factors associated with the onset of MDD (Bernet & Stein, 1999), and a history of CA, is associated not only with this increased prevalence, but also with increased symptom severity and poorer prognosis across a range of mental health disorders (Raposa et al., 2014) and with a lack of response or remission during treatment for depression (Nanni et al., 2012). Whilst CA takes many forms, the pervasive nature of emotional abuse and neglect appears to generate a particular risk factor for mood disorders (Bogdan et al., 2012; Spertus et al., 2003; Watson et al., 2014).

Reduced hippocampal volume is the most consistently documented neural abnormality associated with MDD (Arnone et al., 2016; Campbell et al., 2004; Cole et al., 2011; Han et al., 2016; McKinnon et al., 2009; Roddy et al., 2019; Schmaal et al., 2016; Sheline et al., 1996; Videbech & Ravnkilde, 2004; Wise et al., 2017). Right, and particularly left, hippocampal volume reductions have been found to be robustly associated with duration of untreated depressive episodes (Arnone et al., 2013; Sheline et al., 1996), even in the first episode of depression (Cole et al., 2011; but see McKinnon et al., 2009). In MDD, hippocampal volume loss has been used to track disease severity such as number of episodes, illness duration, and treatment resistance (Sheline et al., 2019), and appears to act as a potential risk factor for new onset depression (Amico et al., 2011). These volume reductions are consistent with the basic science describing the deleterious effects of chronic stress on hippocampal integrity (Magariños & McEwen, 1995), and the observed reductions in hippocampal neurogenesis in rats following chronic restraint stress in adulthood (Pham et al., 2003) or early life (Yun et al., 2010).

CA may alter neural developmental trajectories with structural changes emerging during the transition between puberty and adolescence (Paquola et al., 2017; Teicher & Samson, 2013). Threat and deprivation appear to exert differential effects (McLaughlin et al., 2014). Evidence for an effect of CA on the hippocampus has come from observations across multiple studies demonstrating hippocampal differences in adults who experienced maltreatment in childhood (Dannlowski et al., 2012; Riem et al., 2015). Mirroring MDD, these effects are often more pronounced in the left hippocampus (Frodl et al., 2010; Riem et al., 2015; Schmaal et al., 2016; Stein et al., 1997). Such observations have led to the suggestion that the heightened risk of psychopathology, conferred by the experience of CA, may be at least partially attributable to the enduring consequences of CA on hippocampal development (Riem et al., 2015).

However, the interaction between CA, psychopathology and neural development is difficult to disentangle. Adult studies of CA frequently (Bremner et al., 1997; Calem et al., 2017; Frodl et al., 2010; Opel et al., 2014; Stein et al., 1997; Teicher et al., 2012; Vythilingam et al., 2002), but not invariably (Cohen et al., 2006; Dannlowski et al., 2012; Edmiston et al., 2011; Paquola et al., 2017), include a high prevalence of people with mood and anxiety. Hence, attempting to establish whether CA-related hippocampal damage, precedes, or is a consequence of a subsequent psychopathology, is challenging, especially when the participants in such studies are typically adults who have established psychiatric disorders. Apparently discordant results (De Brito et al., 2013; Frodl et al., 2017) also complicate.

Cognitive studies have also been used to characterise differences in hippocampal-dependent cognition function as a consequence both of MDD (and psychopathology more broadly) and/or CA. Episodic memory (EM) (defined as the recollection of personally experienced events), and autobiographical memory (ABM) (defined as the subgroup of episodic memories that form part of one’s overarching life narrative (Tulving, 1983)), are often considered the quintessential hippocampal-dependent cognitive function (Scoville & Milner, 1957). Multiple studies have now reported a robust link between impaired autobiographical memory and MDD (for meta analysis, see Liu et al., 2013; for review, see Williams et al., 2007), whereby MDD patients’ description of personally experienced past events lack phenomenological richness and specific details. This phenomenon is referred to as Over-general Autobiographical Memory (OGM).

OGM was found to be associated with childhood physical (but not sexual) abuse in adult patients diagnosed with MDD (Griffith et al., 2016). OGM was also observed in female undergraduate students with a history of childhood emotional abuse, an effect that was more pronounced in participants who had not received support for this abuse (Raes et al., 2005). The link between OGM and CA has also been established in studies using multi-informant reports of CA rather than self-report retrospective accounts (Valentino et al., 2009), and in a large population based sample (n=5,792) of participants enrolled in an ongoing longitudinal cohort study (Crane et al., 2014). Moreover, in a meta-analysis which sought to quantify group differences in OGM, Ono and colleagues found a medium effect size of trauma history (e.g. childhood sexual abuse) on OGM, a large effect of MDD, and a very large effect size of OGM among individuals with PTSD (Ono et al., 2016), indicating a complex relationship between OGM and past/current life experiences/psychopathologies.

Alongside EM and ABM, the human hippocampus also supports the simulation of imagined events, e.g. for episodic future thinking (EFT) and scene construction (SC) (Hassabis et al., 2007; Hassabis & Maguire, 2007; Maguire et al., 2016; Maguire & Mullally, 2013; Mullally et al., 2012; Schacter & Addis, 2007). With respect to EFT (defined as the ability to imagine/simulate events that may occur in one’s personal future), deficits have been documented in individual studies and meta-analyses in depression and bipolar disorder (Hallford et al., 2018), and in PTSD (Brown et al., 2014), and schizophrenia (Hallford et al., 2018; Lyons et al., 2016). Reduced specificity of EFT was observed in adult trauma survivors with PTSD, both with and without comorbid depression (Kleim et al., 2014). Interestingly, the adult trauma survivors whose trauma history included childhood trauma exposure demonstrated more over-general EFT than those whose trauma did not include childhood trauma. Trauma (and, in particular, childhood trauma) may thus render individuals ‘stuck’ in that past trauma and lead to difficulty maintaining future orientation (Holman & Silver, 1998; Kleim et al., 2014). Such a ‘stuckness’ has intuitive consequences for the maintenance of, and risk of relapse for, disorders such as MDD.

As discussed above, the human hippocampus also supports scene construction (SC), defined as the mental construction of internal models of the world in the form of spatially coherent scenes (Hassabis et al., 2007; Mullally et al., 2012). Scene construction is impaired in patients with selective, bilateral hippocampal damage and accompanying amnesia, who are unable to construct/imagine spatially coherent fictitious scenes, such as walking along a busy fishing harbour. These deficits are observed despite the patients knowing the individual elements that would be anticipated in specific scenes, and represent a specific impairment in visualising real-world spaces (Mullally et al., 2012). These neuropsychological (and related neuroimaging) findings placed scene processing/construction as a central and core function of the hippocampus. More specifically, the Scene Construction Theory (Hassabis & Maguire, 2007; Maguire et al., 2016; Maguire & Mullally, 2013) posited that an impaired ability to construct scenes compromises related cognitive functions, such as episodic/autobiographical memory and EFT. That is, impaired SC degrades the stage on which episodic/autobiographical memory recollections and personal future simulations can be (re)enacted. From this theoretical perspective, impairment to the core hippocampal function of scene construction may result in EM/ABM and EFT that lack phenomenological richness and detail (Klein et al., 2002; Maguire & Mullally, 2013; Race et al., 2011).

Whilst SC deficits have been observed in patients with schizophrenia (Raffard et al., 2010) and autism (Lind et al., 2014), little is known about the integrity of scene construction processes in depression, and even less in relation to childhood adversity, maltreatment or trauma. Research into mental imagery abnormalities in depression is in its early stages (for review see Holmes et al., 2016), with initial findings suggesting that individuals with mood disorders generate less vivid and compelling positive future imagery, but an increased frequency in distressing intrusive imagery (e.g. Holmes et al., 2008; MacLeod & Byrne, 1996). It has yet to be explored whether such difficulties are driven by a core SC deficit, however, the description of the mental imagery deficits associated with mood disorders does appear to be consistent with a difficulty imagining three-dimensional spatial contexts (Mullally et al., 2012).

If, as proposed by Scene Construction Theory, SC deficits engender EM/ABM and EFT deficits, then understanding the relationship between childhood adversity, mood/wellbeing, and core hippocampal cognitive processes (such as scene construction), may be essential in unpacking this constellation of vulnerability to psychopathology and impaired cognitive processes (such as OGM and over-general future thinking). Here we aimed to explore these hippocampal-dependent cognitive processes in relation to chronic early-life stress in a non-clinical cohort of young adults. We focused on EM as opposed to ABM as we elected to focus our investigations on more emotionally neutral events. We also chose to focus on childhood emotional neglect (CEN) for scientific (Edmiston et al., 2011; Frodl et al., 2010) and pragmatic reasons. We selected, as our population, undergraduate students. We also sought to explore the relationships between current wellbeing and mood, retrospectively reported CEN and the hippocampal-dependent cognitive processes.

It was hypothesised that an inverse relationship would be observed between CEN and current wellbeing/depressive symptomology, between CEN and performance on the hippocampal-dependent cognitive tasks, and between current wellbeing/depressive symptomology and performance on these tasks.

## Methods

### Participants and Procedure

The study incorporated two phases and used two separate cohorts, 2 years apart, and was approved by the Faculty of Medical Sciences Research Ethics Committee, part of Newcastle University’s Research Ethics Committee.

#### Phase 1

An initial online survey was distributed. The survey included (i) the five-item emotional neglect subscale of the Childhood Trauma Questionnaire (CTQ-EN). The CTQ is a validated 28-item self-report questionnaire used to provide a retrospective measure of childhood trauma (Bernstein et al., 2003). The questions are split into five subscales, one of which is emotional neglect (EN). The CTQ-EN items, each start with the stem “When I was growing-up” and are followed by a statement, such as “I felt loved” (Bernstein, 1998). Participants are asked to respond using a 5-point Likert scale ranging from, “never true” to “very often true”. Distributions of these scores across Bernstein and Fink (1998) classification categories can be seen in Table 1 (see also Supplementary Materials Part 1 for histograms of score distributions across Cohorts 1 and 2;). Wellbeing was also measured using the Office for National Statistics’ (ONS) four-item Subjective Wellbeing Survey (Davies, 2014) (see also ONS, 2018). Here, respondents indicate on a scale of 0 (“Not at all”) to 10 (“Extremely”) the response to: Q1: “Overall, how satisfied with your life are you nowadays?”, Q2: “Overall, to what extent do you feel the things you do in your life are worthwhile?”, Q3; “Overall, how happy did you feel yesterday?”, Q4: “Overall, how anxious did you feel yesterday?”. The average score of these four items (with the one negative anxiety item reverse scored) generate a composite wellbeing score for each respondent between 0 (low subjective wellbeing) to 10 (high subjective wellbeing). Standard demographic questions pertaining to age, ethnicity and gender were also included. In cohort 1, the survey was sent electronically to a convenience sample of 9,985 Newcastle University undergraduate students, from whom 1,027 completed responses were collected (313 males, 691 females, 23 undisclosed; mean age = 20.27, SD = 1.9, range = 18-30). For cohort 2, the distribution was to all first-year students based at the University’s Newcastle upon Tyne campus, from which 458 responses were collected (324 females, 129 males, 3 non-binary, 2 undisclosed; mean age=19.2, SD=1.56, range=18-30 years) (see Table 1 for further details). In Cohort 2, additional questions were included in Phase 1; participants were asked to rate aspects of their general memory ability, plus their EFT abilities, using a five-point rating scale (see Table 2 for specific questions and scoring details) using ten questions which correspond with the traditional definitions of episodic memory, semantic memory and episodic future thinking selected from the Survey of Autobiographical Memory (SAM) (Palombo et al., 2013). In addition, participants were asked to reflect on how easily they found it to generate visual mental images (see Table 2) and to bring to mind and reflect on their earliest memories. Responses to the latter question will be reported elsewhere.

**Table 1.** The estimated proportion of the sample belonging to each of Bernstein and Fink’s (1997) EN categories, plus mean wellbeing score (as measured by the ONS four wellbeing measures assessing general life satisfaction, general life worth, and happiness and anxiety of the previous day summed to create an overall composite wellbeing measure (whereby the minimum score = 1 and the maximum score = 40) and standard deviation for each group.

| CTQ-EN Score | Classification | n (%) |  |  | Total Wellbeing Score [mean (SD)] |  |  |
| --- | --- | --- | --- | --- | --- | --- | --- |
|  |  | Cohort 1 (Phase 1) | Cohort 2 (Phase 1) | Combined P1 Cohorts | Cohort 1 (Phase 1) | Cohort 2 (Phase 1) | Combined P1 Cohorts |
| 5 – 9 | None (to minimal) | 631 (61.4%) | 247 (53.9%) | 878 (59.1%) | 29.2 (5.7) | 26.4 (6.4) | 28.4 (6.0) |
| 10 – 14 | Low (to moderate) | 292 (28.4%) | 129 (28.2%) | 421 (28.4%) | 26.3 (6.2) | 23.8 (6.7) | 25.5 (6.5) |
| 15-17 | Moderate (to severe) | 70 (6.8%) | 49 (10.7%) | 119 (8.0%) | 24.9 (6.7) | 23.2 (6.5) | 24.2 (6.7) |
| ≥18 | Severe (to extreme) | 34 (3.3%) | 33 (7.21%) | 67 (4.5%) | 22.7 (8.2) | 21.1 (6.3) | 22.0 (7.3) |
|  |  | Cohort 1 (Phase 2) | Cohort 2 (Phase 2) | Combined P2 Cohorts | Cohort 1 (Phase 2) | Cohort 2 (Phase 2) | Combined Phase 2 Cohorts |
| 5 – 9 | None (to minimal) | 8 (34.8%) | 22 (53.6%) | 30 (46.9%) | 29.8 (4.7) | 26.0 (7.4) | 26.8 (7.0) |
| 10 – 14 | Low (to moderate) | 11 (47.8%) | 6 (14.6%) | 17 (26.6%) | 28.9 (5.6) | 23.4 (9.0) | 26.9 (7.2) |
| 15-17 | Moderate (to severe) | 2 (8.7%) | 5 (12.2%) | 7 (10.9%) | 22.0 (9.9) | 15.1 (7.3) | 17.1 (7.9) |
| ≥18 | Severe (to extreme) | 2 (8.7%) | 8 (19.5%) | 10 (15.6%) | 19.5 (12.0) | 19.5 (8.7) | 19.5 (8.6) |

**Table 2.** **[*Phase 1 (Cohort 2 only)*].** Online questions exploring participants’ subjective judgements of their experiences recollecting their personal pasts (i.e. episodic memory recollection) and their learnt knowledge (i.e. semantic memory), simulating their potential personal futures (i.e. episodic future thinking), and their overall ease of visual mental imagery. Participants were requested to endorse the response most applicable to their experience, from a total of five response options that ranged from totally disagree to totally agree. These were scored as follows: 1 = totally disagree, 2 = disagree somewhat, 3 = Neither agree nor disagree, 4 = Agree somewhat, 5 = totally agree. The ‘High Score’ column indicates whether a high score represents a positive or negative endorsement of the statement. Columns 4-7: Spearman’s correlations and confidence intervals for relationships between CEN and Wellbeing, with the self-report responses to questions about their experience recollecting episodic memories, factual information (i.e. semantic memories), imagining personal future events (EFT), and ease of visual imagery generation. Shaded findings indicate significant results (at 95% CI).

|  |  | High Score | Childhood EN (CTQ) |  |  | Wellbeing (Total) |  |  |
| --- | --- | --- | --- | --- | --- | --- | --- | --- |
|  |  |  | r<sub>s</sub> | Sig. | CI | r<sub>s</sub> | Sig. | CI |
| Episodic Memory |  |  |  |  |  |  |  |  |
| Q1. Detail | “When I remember past events, I remember a lot of details” | Positive | -.050 | .283 | -.142, .039 | .102 | .030 | .007, .193 |
| Q2. Difficulty | “Specific past events are difficult for me to recall” | Negative | .091 | .050 | -.001, 0.183 | -.179 | .001 | -.272, -.089 |
| Q3. Confidence | “I am highly confident in my ability to remember past events” | Positive | -.144 | .002 | -.232, -.057 | .211 | .001 | .123, .296 |
| Q4. Event Order | “When I remember past events, I have a hard time determining the order of details in the event” | Negative | .026 | .567 | -.062, .119 | -.092 | .050 | -.181, -.003 |
| Semantic Memory |  |  |  |  |  |  |  |  |
| Q1. Fact Difficulty | “I can learn and repeat facts easily, even if I don’t remember where I learned them” | Positive | -.041 | .384 | -.131, .049 | .113 | .015 | .022, .205 |
| Q2. Information Difficulty | “I have a hard time remembering information I have learned at school, university or work” | Negative | .143 | .002 | .049, .233 | -.240 | .001 | -.327, -.153 |
| Q3. General Info Difficulty | “I am very good at remembering information about people that I know e.g., the names of other people’s friends/family members etc.” | Positive | -.053 | .254 | -.141, .036 | .114 | .015 | .026, .200 |
| Episodic Future Thinking |  |  |  |  |  |  |  |  |
| Q1. Difficulty | “I have a difficult time imagining specific events in the future” | Negative | .110 | .018 | .024, .196 | -.274 | .001 | -.358, -.181 |
| Q2. Emotions | “When I imagine an event in the future, I can imagine how I may feel” | Positive | -.094 | .045 | -.178, -.010 | .176 | .001 | .087, .263 |
| Q3 Specificity | “When I imagine an event in the future, the event generates vivid mental images that are specific in time and place” | Positive | .068 | .148 | -.032, .155 | .109 | .020 | .011, .207 |
| Visual Imagery |  |  |  |  |  |  |  |  |
| Q1. Difficulty | “I find it easy to visualise images or pictures in my mind’s eye” | Positive | -.106 | .023 | -.202, -.005 | .139 | .003 | .047, .232 |

#### Phase 2

A subset of phase 1 participants were invited to take part in a second experimental testing session. For cohort 1, in order to generate a sample with a spread of CEN scores, cluster analysis was used, which generated three categories, characterized as low (n = 544, 52.6%), moderate (n = 280, 27.1%), and high (n = 209, 20.2%) childhood adversity. Random selection within each category generated a sample of 32 who were invited to participant in Phase 2. Of these, 27 agreed to complete Phase 2 testing (12 male, 15 female), four of these were excluded from phase 2 because they were second-language English speakers. Of the remaining twenty-three participants (11 male, 12 female; mean age = 20.87, SD = 2.4, range: 18-28; CTQ-EN: *M* = 11.61, SD 4.30, range: 5-21), using Bernstein and Fink’s 2003 categories, 34.78% were categorised as ‘none (to minimal)’, 47.83% as ‘low (to moderate)’, 8.7% as ‘moderate (to severe)’, and 8.7% as ‘severe (to extreme)’ (see Table 1 for further details). The smaller size of Cohort 2 suggested a slightly different approach for sample selection. Here all participants who fell within Bernstein and Fink’s ‘moderate (to severe)’ or ‘severe (to extreme)’ groups, and for whom English was a first language, were invited to take part in Phase 2 testing (n = 70). For each participant who accepted this invitation, a participant, matched for gender and wellbeing, from the ‘none (to minimal)’ or ‘low (to moderate)’ groups, was also invited to participate in Phase 2 testing. This matched recruitment strategy was subsequently abandoned due to very low uptake of the invitation and was replaced with a more general blanket invitation to approximately 70 from the ‘none (to minimal)’ or ‘low (to moderate)’ groups. In total forty-one participants completed this session (14 male, 27 female; Mean Age = 19.75; SD = 2.44; range: 18-30; CTQ-EN: *M* = 11.07, SD 5.63, range: 5-21), 53.66% were categorised as ‘none (to minimal)’, 14.63% as ‘low (to moderate)’, 12.19% as ‘moderate (to severe)’, and 19.51% as ‘severe (to extreme)’ (see Table 1 for further details).

In phase 2, episodic memory, episodic future thinking, scene construction (Cohorts 1 and 2), and anterograde memory (Cohort 2 only) were assessed. Identical instructions and procedures for EFT, EM and SC were used in both cohorts; however, participants in Cohort 2 performed a reduced number of trials for each of these measures. Specifically, as opposed to recollecting (EM) or imaging (EFT) events that occurred/would occur one week, one month, one year and five years ago/in the future (as in Cohort 1), participants in Cohort 2 only recollected and imagined events that occurred/would occur one month and five years ago/in the future. Similarly, whilst Cohort 1 participants imagined three fictitious scenes, Cohort 2 participants only imagined two (see Figure 1 for further details). A second measure of self-reported CEN was collected at phase 2. The mean CTQ-EN scores from Phases 1 and 2 was used in analysis because we assume an aggregate measure of CEN obtained at two separate time points is a more accurate estimate of CEN. There was a strong correlation between phase 1 and phase 2 CEN scores (r_s_=0.923, [0.815, 0.970]), indicating a good degree of consistency. At phase 2, additional measures of wellbeing and depression, were also administered (see Figure 1 for further details). Both Cohort 1 and Cohort 2 participants also completed the Beck Depression Inventory-Second Edition (BDI-II) (Beck et al., 1996) and the State-Trait Anxiety Inventory (STAI) (Spielberger et al., 1983). The BDI-II is a widely used self-report measure of depressive symptoms, with a two-factor structure measuring cognitive-affective and somatic depressive symptoms. It is used in both clinical and non-clinical populations, and the validity and reliability of its use in college students has been supported by multiple empirical studies (e.g. Storch et al., 2004). It has also been shown to be acceptable as a screening tool for major depressive episode in a sample of healthy adults, (Kjaergaard et al., 2014). The STAI is a widely used self-report measure of trait- and state-anxiety. In college students, the trait-anxiety component has been shown to be a reliable measure of the student’s individual level of anxiety across a test-retest delay, whilst the state-anxiety component has been found to discriminate between transient high-stress (i.e. post-examination) and low-stress situations (Metzger, 1976).

**Figure 1:**
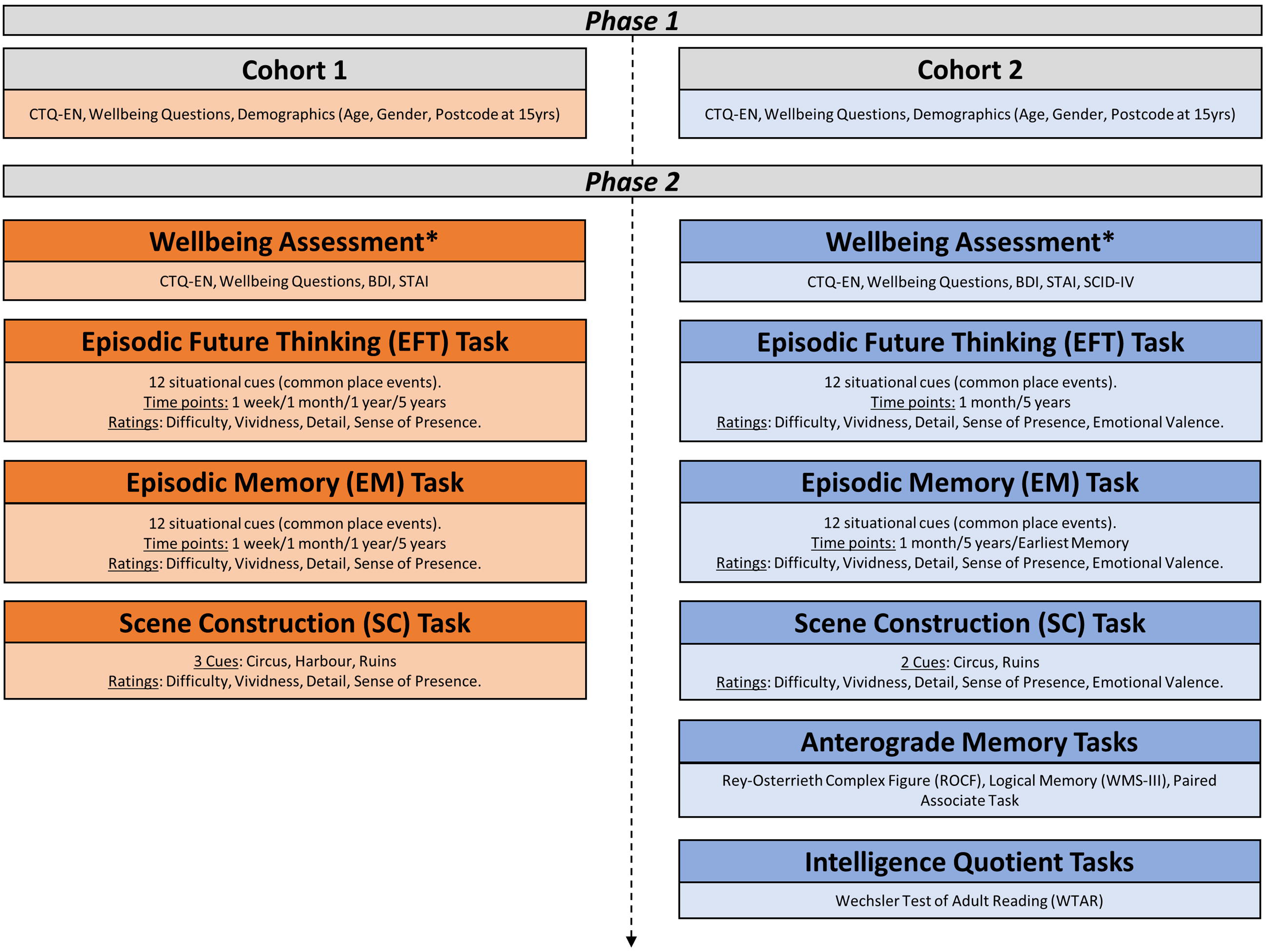
Flowchart of tasks in both Phase 1 and Phase 2 in Cohort 1 (2014/2015) and Cohort 2 (2016/2017). CTQ-EN: Emotional Neglect subscale of the Childhood Trauma Questionnaire (Bernstein et al., 2003). BDI: Beck Depression Inventory (Beck et al., 1996). STAI: State Trait Anxiety Inventory (Spielberger et al., 1983). SCID-IV: Structured Clinical Interview for DSM-IV (First et al., 2015). The Rey-Osterrieth Complex Figure task (ROCF) (Rey & Osterrieth, 1993). WMS-III: The Wechsler Memory Scale, 3^rd^ Edition, Logical Stories subtest (Wechsler, 1997). Paired Associates task (Anderson et al., 2004; Benoit & Anderson, 2012). Weschler Test of Adult Reading (WTAR) (Weschler, 2001).

At phase 2, cohort 2 participants also performed additional cognitive assessments; specifically the Weschler Test of Adult Reading (WTAR) (Weschler, 2001), Weschler Memory Scale III, Logical Stories subtest (Wechsler, 1997), the Rey-Osterrieth Complex Figure task (ROCF) (Rey & Osterrieth, 1993), and the Paired Associates Task (see Figure 1). The WTAR provides a rapid reading-based IQ-estimate. It was developed and co-normed with the Weschler Adult Intelligence Scale-III in a representative sample of normally functioning adults, enabling the conversion of WTAR scores to Full Scale IQ (FSIQ) estimates. Whilst it is widely used as a measure of pre-morbid IQ, it was included in this assessment battery as a control measure to monitor for any variation in estimated-IQ across the CEN spectrum. The Logical Stories and ROCF were included as both represent established and widely used measures of verbal and visual memory respectively. Deficits in episodic memory are the most consistently reported neuropsychological deficits following hippocampal-damage, and these deficits are traditionally assessed using standardised tasks of anterograde memory. In a review of 147 amnesic patients, consistent deficits were noted on tasks of diagram recall (e.g. the ROCF), and paired associate, list, and story recall (along with more inconsistent impairment on tests of verbal and visual recognition) (Spiers et al., 2001). As standardised paired associate tests tend to show ceiling effects when used in undergraduate populations, we devised a more challenging measure of list learning based on a materials developed for use in a similar population (Anderson et al., 2004; Benoit & Anderson, 2012) (see Supplementary Material Part 2). Participants in both cohorts were also asked to report their postcode at age 15 in order to calculate Multiple Deprivation Index scores, however this data was not analysed in the current study and will be reported elsewhere.

### Neuropsychological Tasks

For both EFT and EM tasks, participants were instructed to read through a list of twelve commonplace event situational cues (e.g. going on a shopping trip/eating out in a restaurant/going for a walk in a forest/attending a wedding) and decide if they could realistically imagine themselves in any of these scenarios in one month’s time/one month ago, one year’s time/one year ago, and in five years’ time/five years ago. Participants were only permitted to use a single cue once. If participants could not realistically envision or recollect any of these situations occurring within the given timeframes, they were permitted to self-generate a cue. This option was not chosen by any of the participants. Participants were also instructed that the imagined future event/recalled event should last between a few minutes or hours but it should not last a whole day, nor be a highly significant emotional event and that the event they recalled/imagined needed to be specific in time and place. Once cues were selected, the experimenter read the cue aloud alongside its temporal label (e.g. “in five years’ time when you imagine that you will be attending a wedding”/”five years ago when you attended a wedding”.). Temporal order was counter-balanced across participants. Participants then described the event aloud. If the description was deemed brief and/or lacking in significant detail, the experimenter probed with, “Is there anything else you can tell me about this event?”. At the end of each trial (unless all aspects of this question had already been given in detail by the participant), all participants were asked “Can you tell me any more about where and when this event is taking place, who is there, how you feel, and what you are thinking?”. For the SC task, participants were required to visualise and describe three fictitious scenes: a circus, a harbour and a derelict building (e.g. “imagine that you are exploring the ruins of a derelict building”). Detailed and explicit instructions were provided to participants requesting that they picture the whole scene in their mind’s eye, as vividly and in as much detail as possible, and to use all available senses (such as sight, sound and smell) whilst doing so. In order to reinforce these instructions, an example of a vivid SC (based on a cue that was not used in participant trials) was provided to participants prior to their first trial, along with the clear instruction to be as vivid and as imaginative as possible. In the SC and EFT trails participants were reminded, prior to commencing each trial, that this was not a memory task and therefore that they should not simply imagine and describe a memory of a similar place.

Participants completed a number of self-evaluation questions pertaining to their experience of visualising scenes or episodes (see Table 3 for details). Participants then completed the spatial coherence index (SCI) questionnaire that contains twelve statements requiring a true or false response. Eight of these statements depict a spatially contiguous scene (e.g. “The scene that I visualised when imagining this event was similar to looking at a picture of it or seeing it on TV”), and four depict a spatially fragmented scene (“What I visualised wasn’t a scene you could step into”). The combination of statements endorsed are used to generate the SCI (a measure of the imagined/remembered scenes). Finally, in keeping with previous scoring protocols, an experiential index (EI) score was generated from a combination of the SCI, self-reported vividness and sense of presence (SoP), experimenter ratings of the overall quality of the future simulation/memory/scene, and the number of entities present, spatial references, sensory details, and thoughts, emotions and actions described by participants in their narratives of their recollections/simulations (for full scoring details see Hassabis et al., 2007). As such, the EI acts as a proxy measure of overall richness of the recollection/simulation. (Note: in Cohort 2, both the experimenters and scorers were blind to the CEN score of participants. This was not the case in Cohort 1).

**Table 3.** Self-evaluation questions following each EFT, EM and SC task in Phase 2 (laboratory phase). *Cohort 2 only

| Question | Response ranges (5-point Likert Scale) |  |
| --- | --- | --- |
| “How difficult did you find the task?” | 1 = Very easy | 5 = Very hard |
| “How vivid was the event you imagined in your mind’s eye?” | 1 = Not at all vivid | 5 = Extremely vivid |
| “How detailed was the event you imagined in your mind’s eye?” | 1 = Hardly any details at all | 5 = Extremely richly detailed |
| “How much of a sense of being there did you have when imagining this future event?” | 1 = It didn’t feel like I was there at all | 5 = It strongly felt like I was really there |
| “How much emotion did you experience when imagining this future event?”* | 1 = I felt totally detached | 5 = It was highly emotional |

The EFT assessment was conducted first to avoid carryover effects from either the EM or the SC protocols. SC was assessed last as this was the only one of the three measurement types in which participants were provided with specific instructions to visualise a vivid scene in their mind’s eye, using as much detail as possible, whereas EFT and EM assessments were intended to measure how participants generated these scenes in their future thinking and episodic recollections naturally.

### Data Analysis

All data was analysed using SPSS v.23 (IBM Corp., 2015). Normality assessments using histograms, box-plots, and Shapiro-Wilk and Kolmogorov-Smirnov tests were carried out in order to determine whether analysis would be parametric or non-parametric. Correlation analyses were used to establish relationships between variables (Spearman’s rho when residuals of relationships were not normally distributed) with results reported as test statistics, p-values and confidence intervals. All confidence intervals were set at a 95% threshold. When data were categorised according to CEN severity categories, differences between groups were calculated using ANOVA and Tukey HSD bootstrapping for multiple comparisons. In all analyses, the data was analysed using the pooled cohorts (Cohort 1+2), and in each cohort independently (Cohort 1 and Cohort 2). This was done to provide an internal replication of the findings.

Mediation analyses were carried out for variables that correlated with both CEN and Total Wellbeing (Phase 1) or BDI-II (Phase 2) using the SPSS macro (Hayes, 2018). Seed values were determined randomly on a six-digit magnitude and all confidence intervals were set at a 95% threshold.

## Results

### Phase 1 (Cohorts 1 and 2)

Spearman’s correlation analyses revealed a significant negative relationship between childhood emotional neglect and overall wellbeing (Cohort 1: n=1027, r_s_=-0.308, p<0.001 [-0.368, -0.252], Cohort 2: n=457, r_s_=-0.289, p<0.001 [-0.369, -0.205], Cohorts combined: n=1484, r_s_=-0.308, p<0.001 [-0.353, -0.256]). When both cohorts were analysed separately, and when the two cohorts were combined, CEN significantly correlated with all four of the wellbeing questions, except for anxiety in Cohort 2 (see Figure 2 and Supplementary Materials Part 3 for further details).

**Figure 2.**
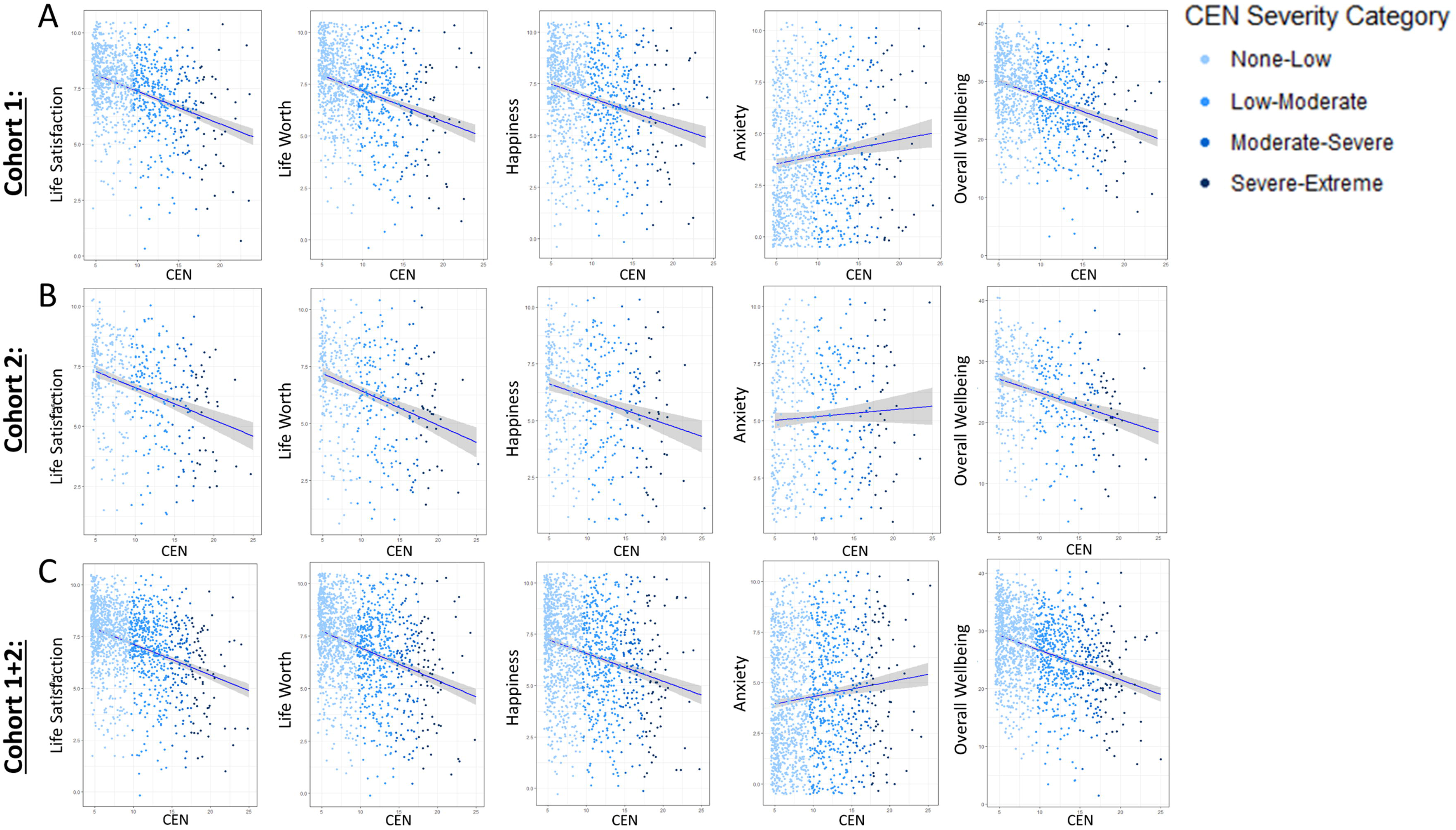
Phase 1 (Cohorts 1 and 2). Jitter plots with regression lines of the relationships between CEN and Overall Wellbeing, Life Satisfaction, Life Worth, Happiness and Anxiety for cohort 1 (Panel A), cohort 2 (Panel B) and the phase 1 cohorts combined (Panel C). Horizontal and vertical jitter = .5. (Note: For the Overall Wellbeing score, Anxiety is reverse scored before being summed with the three other components).

In order to explore the comparability of our samples to previously described samples we firstly looked at previously published UK National Averages from the public mental health survey (Davies, 2014). This demonstrated that compared to UK National average levels, “Satisfied”, “Worthwhile” and “Happiness” were rated as lower by our participants, whilst “Anxiety” was higher (see Figure 3a and Supplementary Materials Part 4 for further details). The difference between each of the four measures and the national averages was more pronounced in Cohort 2. In order to illustrate the negative association between CEN and wellbeing in a manner consistent with previous reports, Phase 1 participants (in both cohorts) were categorised by degree of emotional neglect according to the CTQ divisions of “none”, “low”, “moderate” and “severe”, and responses to each of the four ONS wellbeing questions probed further (see Figure 3b and Supplementary Materials Part 5 for further details). Overall, these analyses found that there were significant between-group differences for Life Satisfaction scores; for Worthwhile and Happiness, between-group differences generally occurred between the none/low groups and higher CEN groups, than between higher severity categories such as between Moderate and Severe; and no significant between-group differences were observed for Anxiety scores. These findings were generally well replicated in Cohort 2, with significant differences in mean scores between CEN severity categories being found in all measures but anxiety, with significant differences occurring more frequently between the none/low scoring groups and the severe scorers, than between moderate and severe scorers (see Figure 3b).

**Figure 3.**
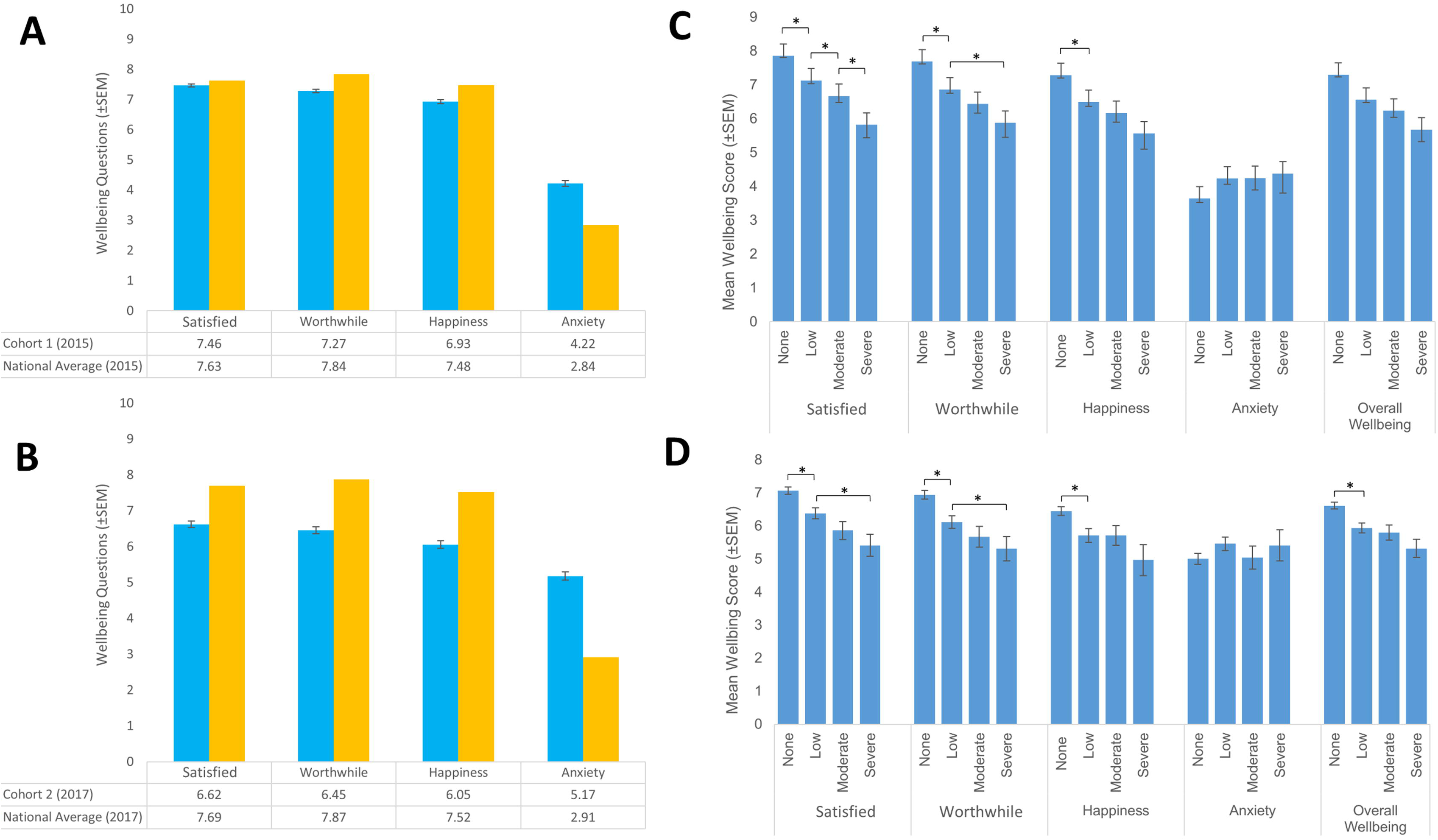
Phase 1. <u>Panel A (Cohort 1) and Panel B (Cohort 2)</u>: Mean Phase 1 scores of the four ONS Wellbeing questions alongside national average scores for the same year (Personal Wellbeing Estimates: July 2014 – June 2015 and January 2017 – December 2017, Office for National Statistics, 2018). <u>Panels C (Cohort 1) and Panel D (Cohort 2)</u>: Phase 1 participants were categorised based on their CEN scores into the CTQ divisions: None (5-9), Low (10-14), Moderate (15-17) and Severe (18-25) categories (full range of scores: 5-25). Asterisks denote significant differences in Wellbeing question scores between CEN categories as revealed by Tukey HSD Bootstrapping for Multiple Comparisons at 95% confidence intervals.

### Phase 1 (Cohort 2 only): Self-Reported Measures of Memory, Future Thinking, and Mental Imagery

As evident in Table 2, when participants (n=456) rated their own memory (for both episodic and semantic information, their experiences of imagining their personal futures (EFT), and the ease of their mental imagery (MI), small negative correlations were observed between self-reported levels of CEN and self-reported levels of difficulty recollecting specific episodic memories (EM Q2), CEN and episodic memory confidence (EM Q3), CEN and memory for information learnt at school/university/work (Semantic Memory Q2), CEN and difficulty simulating future events (EFT Q1), CEN and difficulty imagining emotions in future events (EFT Q2), and ease of visual imagery (Visual Imagery Q1). In all instances, higher CEN related to poorer scores on each parameter. Similar small, but positive, correlations were observed between self-reported levels of total wellbeing and each of these variables (i.e. EM, SM, EFT and MI); whereby lower levels of total wellbeing related to lower ratings on each parameter.

#### Mediation Analyses

Given this multitude of correlations with both current Wellbeing, and to a lesser extent ratings of childhood emotional neglect (CEN), we further explored the relationships between variables that correlated significantly with both CEN and current wellbeing (i.e. Episodic Memory Q3, Semantic Memory Q2, EFT Q1 and Q2, and Visual Imagery) using mediation analyses (Preacher & Hayes, 2004). This enabled us to ask whether the predictor variable (CEN) had a causal effect on the dependent variable of interest (e.g. Episodic Memory Q3) via a third, mediator variable (current Wellbeing). More specifically, we asked whether current Wellbeing mediated the relationship between CEN and difficulty recollecting specific episodic memory (EM Q2), episodic memory confidence (EM Q3), CEN and memory for information learnt at school/university/work (Semantic Memory Q2), CEN and difficulty simulating future events (EFT Q1), CEN and difficulty imagining emotions in future events (EFT Q2), and ease of visual imagery (Visual Imagery Q1). The SPSS macro PROCESS v3.5 (Hayes, 2018) was used to perform simple mediation analyses with Total Wellbeing as a mediator. We found significant effects of the mediation pathway (using 5000 bootstraps samples, 95% CI) in five of the six analyses i.e., there were significant indirect effects of current Wellbeing on the relationship between 1) CEN and difficulty recollecting specific episodic memory (EM Q2: Indirect effect = 0.0130, CI [0.0400, 0.0229]; Figure 4A), 2) CEN and confidence in recollected episodic memories (EM Q3: Indirect effect = -0.0152, CI [-0.0250, -0.0067]; Figure 4B), 3) CEN and Semantic Memory Q2 (Indirect effect = 0.0213, CI [0.0114, 0.0329, Figure 4C]), 4) CEN and EFT difficulty (EFT Q1: Indirect effect = 0.0248, CI [0.0148, 0.0359], Figure 4D), and between 5) CEN and EFT emotions (EFT Q2: Indirect effect = -0.0174, CI [-0.0280, -0.0085], Figure 4E). This demonstrates that for the relationships between CEN, current wellbeing and each of these individual dependent variables were not independent. However, no significant effect of the mediation pathway was observed in the sixth relationship explored (Figure 4F); where we asked whether CEN had a causal effect on the ease of Visual Imagery via current Wellbeing (Indirect effect = -0.0077, CI [-0.0173, 0.0006]). This demonstrates that the relationships between CEN and ease of Visual Imagery, and between current Wellbeing and ease of Visual Imagery, may occur independently of one another.

**Figure 4:**
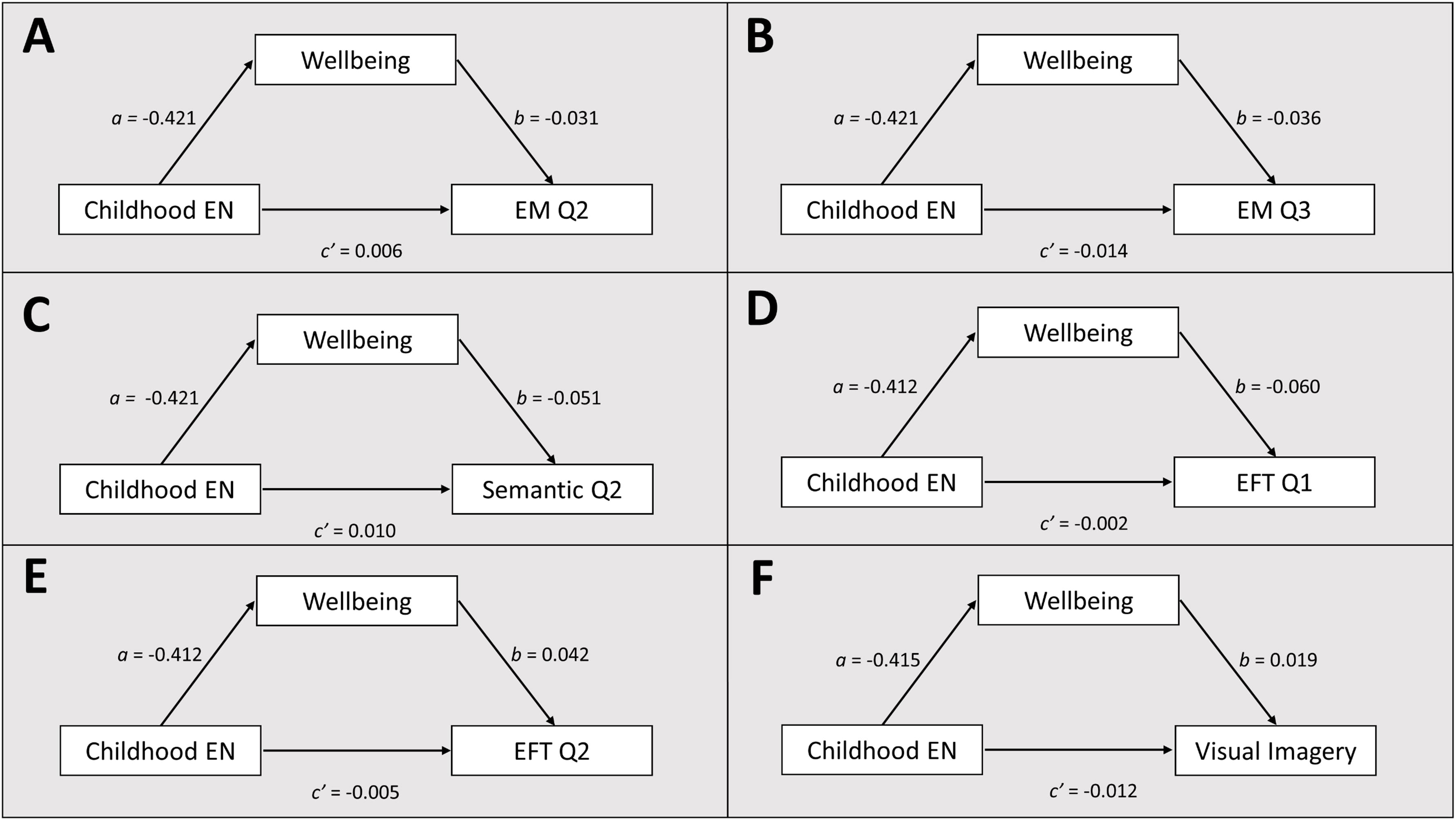
Phase 1: Mediation Analyses (Cohort 2). <u>Panel A</u>: Mediation analysis concluded that current Wellbeing mediated the relationship between CEN and participants’ self-reported level of difficulty recollecting specific past events (Episodic Memory Q2). <u>Panel B</u>: Mediation analysis concluded that current Wellbeing mediated the relationship between CEN and participants’ confidence in their abilities to remember past events (Episodic Memory Q3). <u>Panel C</u>: Mediation analysis concluded that current Wellbeing mediated the relationship between CEN and participants’ self-reported difficulties remembering information learned at school, university or work (Semantic Memory Q2). <u>Panel D</u>: Mediation analysis concluded that current Wellbeing mediated the relationship between CEN and difficulty imagining specific future events (EFT Q1). <u>Panel E</u>: Mediation analysis concluded that current Wellbeing mediated the relationship between CEN and participants’ self-reported ability to imagine how they feel when imagining future events (EFT Q2). <u>Panel F</u>: Mediation analysis concluded that total current Wellbeing did *not* mediate the relationship between CEN and ease of visual mental imagery (Visual Imagery Q1). Each plot reports unstandardized regression coefficients for the relationship between the independent variable (CEN) and the dependent variable as mediated by participant ratings of current wellbeing. Significant indirect effect are highlighted in bold.

### Phase 2 (Cohorts 1 and 2) CEN and Depression

Correlations were observed in Phase 2 between CEN and scores on the Beck Depression Inventory (BDI-II) for the combined cohorts (r_s_=0.283, p < 0.05 [0.017, 0.508]). A significant correlation was also observed in Cohort 1 (r_s_=0.419, p < 0.05 [-0.036, 0.741]), but this correlation was not significant at the 0.05 level in Cohort 2 (r_s_=0.278, p = 0.082 [-0.093, 0.582]).

### Phase 2 (Cohorts 1 and 2) CEN and Anxiety

Correlations were observed in Phase 2 between CEN and scores on STAI (Trait Anxiety) for the combined cohorts (r_s_=0.262, p < 0.05 [-0.014, 0.479]) but not between CEN and scores on STAI (State Anxiety) for the combined cohorts (r_s_=0.179, p = 0.163 [-0.089, 0.410]). In Cohort 1 alone, no significant correlations were found Cohort 1 (CEN and Trait Anxiety, r_s_=0.38, p = 0.074 [-0.063, 0.708]; CEN and State Anxiety, r_s_=0.357, p = 0.094 [-0.087, 0.755]), although these same correlations were significant in Cohort 2 (CEN and Trait Anxiety, r_s_=0.35, p < 0.05 [-0.016, 0.629]; CEN and State Anxiety, _rs_=0.369, p < 0.05 [0.012, 0.666]).

### Phase 2 (Cohorts 1 and 2) Depression, Anxiety and Wellbeing

Strong correlations were observed between depression scores (BDI-II) and both STAI trait (r_s_=0.874, p < 0.001 [0.737, 0.905]) and state (r_s_=0.771, p < 0.001 [0.613, 0.859]) anxiety. Similarly, strong correlations were also observed between overall estimates of wellbeing and both STAI trait (r_s_=-0.639, p < 0.001 [-0.779, -0.434]) and STAI state (r_s_=-0.564, p < 0.001 [-0.712, -0.349]) anxiety. Estimates of wellbeing and BDI scores also correlated (r_s_=-0.658, p < 0.001 [-0.788, -0.463]).

### Phase 2 (Cohorts 1 and 2): Performance on laboratory-based tasks of episodic memory, episodic future thinking and scene construction

In the second phase of study, participants’ subjective experiences whilst imagining/recollecting EFTs, scenes, and EMs were assessed. In Cohort 1, EI scores (which index the richness of these experiences) for EFT, EM and SC (and for the composite measure ‘EFT+EM+SC’) correlated negatively with CEN, such that higher CEN related to lower EI scores. These correlations were not seen in Cohort 2, but were evident in the data from the two cohorts was combined (Cohorts 1+2) for the SC and ‘EFT+EM+SC’ data. On the other hand, whilst BDI-II scores correlated with EI the SC and ‘EFT+EM+SC’ in the Cohort 1 dataset, this not were not replicated in Cohort 2 nor did the correlations remain significant when the data from two cohorts (Cohorts 1+2) were combined (although for both SC and EFT+EM+SC the p-values for the combined cohorts were < 0.1; see Table 4).

**Table 4.** Phase 2: Spearman’s correlations and confidence intervals for relationships between CEN and BDI, and EFT/EM/SC variables for Cohorts 1 and 2, as well as the two cohorts combined. Shaded findings indicate significant results (at p < 0.05; lighter shading indicates p < 0.1).

|  |  | CEN |  |  |  |  |  |  |  |  | BDI |  |  |  |  |  |  |  |  |
| --- | --- | --- | --- | --- | --- | --- | --- | --- | --- | --- | --- | --- | --- | --- | --- | --- | --- | --- | --- |
|  |  | Cohort 1 |  |  | Cohort 2 |  |  | Cohorts 1+2 |  |  | Cohort 1 |  |  | Cohort 2 |  |  | Cohorts 1+2 |  |  |
|  |  | r<sub>s</sub> | Sig. | CI | r<sub>s</sub> | Sig. | CI | r<sub>s</sub> | Sig. | CI | r<sub>s</sub> | Sig. | CI | r<sub>s</sub> | Sig. | CI | r<sub>s</sub> | Sig. | CI |
| Episodic Future Thinking | EI | -.780 | .000 | -.89, -.53 | .064 | .695 | -.24, .36 | -.225 | .076 | -.45, .03 | -.398 | .060 | -.68, -.02 | -.066 | .691 | -.39, .26 | -.183 | .155 | -.42, .06 |
|  | SCI | -.229 | .293 | -.67, .21 | -.170 | .295 | -.45, .18 | -.163 | .202 | -.39, .09 | -.192 | .379 | -.56, .26 | -.080 | .627 | -.43, .28 | -.110 | .394 | -.37, .16 |
|  | Difficulty | .105 | .634 | -.37, .57 | .229 | .155 | -.77, .50 | .168 | .187 | -.09, .41 | -.105 | .633 | -.37, .58 | .020 | .906 | -.29, .34 | .020 | .878 | -.23, .29 |
|  | Vividness | -.496 | .016 | -.73, -.12 | -.313 | .049 | -.56, -.01 | -.324 | .009 | -.53, -.13 | -.351 | .100 | -.67, .07 | -.070 | .670 | -.38, .27 | -.137 | .288 | -.38, .10 |
|  | Detail | -.434 | .039 | -.74, -.01 | -.244 | .129 | -.46, .02 | -.280 | .026 | -.47, -.06 | -.242 | .266 | -.63, .20 | -.065 | .694 | -.36, .25 | -.083 | .519 | -.33, .16 |
|  | SoP | -.444 | .034 | -.72, -.03 | -.299 | .061 | -.54, -.01 | -.330 | .008 | -.53, -.10 | -.439 | .036 | -.75, -.05 | -.084 | .610 | -.38, .24 | -.192 | .136 | -.43, .06 |
|  | Emotion |  |  |  | -.342 | .042 | -.56, -.02 |  |  |  |  |  |  | -.066 | .688 | -.40, .31 |  |  |  |
| Episodic Memory | EI | -.783 | .000 | -.88, -.57 | .095 | .554 | -.27, .42 | -.146 | .248 | -.39, .13 | -.335 | .118 | -.67, .10 | -.076 | .642 | -.40, .26 | -.178 | .162 | -.42, .10 |
|  | SCI | .256 | .238 | -.16, .67 | -.203 | .203 | -.46, .07 | -.067 | .598 | -.30, .17 | .015 | .947 | -.37, .45 | -.136 | .403 | -.44, .20 | -.044 | .731 | -.28, .22 |
|  | Difficulty | -.241 | .269 | -.64, .20 | -.125 | .436 | -.46, .17 | -.110 | .385 | -.36, .16 | .082 | .709 | -.40, .47 | .125 | .443 | -.20, .43 | .005 | .970 | -.24, .26 |
|  | Vividness | -.032 | .884 | -.51, .42 | -.272 | .085 | -.51, .03 | -.196 | .121 | -.44, .04 | -.254 | .242 | -.57, .17 | -.126 | .438 | -.43, .22 | -.086 | .501 | -.31, .14 |
|  | Detail | .138 | .531 | -.35, .49 | -.057 | .725 | -.32, .24 | -.031 | .811 | -.27, .21 | -.166 | .449 | -.50, .25 | -.165 | .310 | -.49, .17 | -.088 | .492 | -.33, .16 |
|  | SoP | -.117 | .596 | -.55, .29 | -.128 | .426 | -.40, .17 | -.145 | .354 | -.36, .10 | -.236 | .279 | -.61, .18 | .031 | .851 | -.31, .37 | -.027 | .833 | -.28, .23 |
|  | Emotion |  |  |  | -.307 | .051 | -.59, .02 |  |  |  |  |  |  | -.208 | .197 | -.52, .15 |  |  |  |
| Scene Construction | EI | -.753 | .000 | -.92, -.46 | -.232 | .144 | -.53, .10 | -.355 | .004 | -.57, -.11 | -.494 | .017 | -.78, -.09 | -.072 | .658 | -.39, .24 | -.230 | .070 | -.46, .02 |
|  | SCI | -.101 | .646 | -.50, .34 | -.241 | .130 | -.55, .10 | -.192 | .128 | -.44, .06 | -.078 | .723 | -.51, .31 | -.154 | .341 | -.47, .17 | -.106 | .409 | -.35, .15 |
|  | Difficulty | .556 | .006 | .20, .79 | .262 | .099 | -.07, .56 | .311 | .012 | .05, .54 | .360 | .091 | -.10, .75 | .046 | .777 | -.31, .38 | .142 | .268 | -.11, .41 |
|  | Vividness | -.485 | .019 | -.77, -.07 | -.349 | .025 | -.61, -.03 | -.383 | .002 | -.58, -.14 | -.347 | .105 | -.72, .11 | .097 | .551 | -.25, .40 | -.039 | .763 | -.31, .21 |
|  | Detail | -.569 | .005 | -.81, -.23 | -.233 | .142 | -.52, .10 | -.289 | .021 | -.53, -.04 | -.340 | .113 | -.72, 0.12 | .049 | .763 | -.30, .38 | -.067 | .604 | -.34, .18 |
|  | SoP | -.541 | .008 | -.80, -.16 | -.377 | .015 | -.60, -.10 | -.401 | .001 | -.59, -.20 | -.503 | .014 | -.79, -.04 | -.215 | .182 | -.56, .14 | -.295 | .019 | -.55, -.03 |
|  | Emotion |  |  |  | -.461 | .002 | -.65, -.22 |  |  |  |  |  |  | -.045 | .782 | -.33, .29 |  |  |  |
| Combined: EFT+EM+SC | EI | -.838 | .000 | -.93, -.63 | -.077 | .631 | -.36, .25 | -.308 | .014 | -.52, -.04 | -.493 | .017 | -.76, -.10 | -.104 | .523 | -.39, .24 | -.237 | .061 | -.45, .01 |
|  | SCI | -.047 | .831 | -.50, .39 | -.214 | .179 | -.49, .11 | -.163 | .201 | -.41, .11 | -.108 | .624 | -.51, .39 | -.169 | .298 | -.48, .22 | -.105 | .411 | -.38, .16 |
|  | Difficulty | .235 | .281 | -.28, .67 | .210 | .187 | -.13, .47 | .218 | .086 | -.05, .46 | .347 | .105 | -.13, .70 | .092 | .573 | -.20, .37 | .123 | .338 | -.14, .36 |
|  | Vividness | -.499 | .015 | -.75, -.14 | -.385 | .013 | -.60, -.09 | -.423 | .001 | -.60, -.21 | -.512 | .013 | -.83, -.07 | -.035 | .830 | -.34, .31 | -.122 | .340 | -.38, .14 |
|  | Detail | -.434 | .038 | -.76, -.02 | -.218 | .170 | -.47, .07 | -.251 | .047 | -.48, -.02 | -.423 | .045 | -.77, -.01 | -.091 | .575 | -.39, .25 | -.121 | .344 | -.35, .13 |
|  | SoP | -.459 | .028 | -.78, -.09 | -.325 | .038 | -.56, -.02 | -.361 | .004 | -.56, -.12 | -.453 | .030 | -.81, -.02 | -.120 | .462 | -.44, .26 | -.219 | .085 | -.46, .04 |
|  | Emotion |  |  |  | -.431 | .005 | -.68, -.12 |  |  |  |  |  |  | -.154 | .344 | -.46, .19 |  |  |  |

The spatial contiguousness (i.e. the SCI) of the participants’ simulations/recollections did not correlate with either CEN or BDI-II scores in Cohort 1, Cohort 2 or Cohort 1+2 for either of the tasks singularly, or for their combined scores. The lack of a significant relationship between SCI and CEN was surprising in Cohort 1 for a number of reasons: (1) the large effect size noted between the EI and CEN; (2) the fact that the SCI score accounts for approximately 10% of the EI score itself (for more details, see Hassabis et al., 2007); and (3) the tight coupling between these two performance measures noted in previous patient groups (e.g. Hassabis et al., 2007; Mullally et al., 2012).

In order to gain traction over the precision of our estimates, we constructed visual representations on each point and interval estimate for the CEN and EI, and the CEN and SCI analyses, for both the individual and the combined cohorts (see Figure 5 Panels A-B). For the SCI data (Panel B), the 95% confidence intervals surrounding the point estimates in Cohort 1 are wider than in Cohort 2; indicating an increase of precision in the larger of the two samples (i.e. in Cohort 2). Additional studies, with larger sample sizes are required to further explore the relationship between CEN and SCI further.

**Figure 5.**
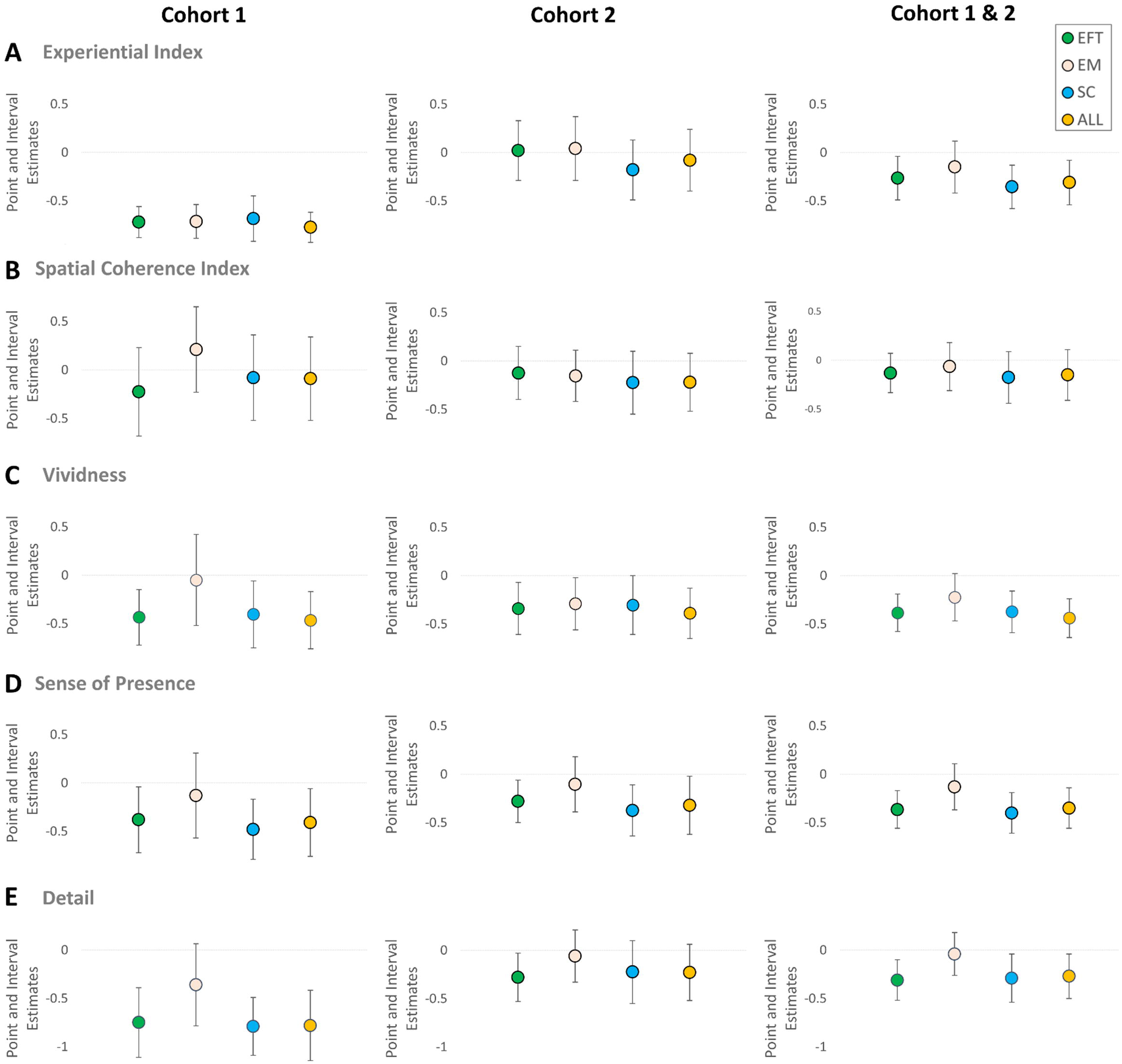
Panels A-E depict the visual representations of point and interval estimate for the CEN and ‘Experiential Index’ (EI) analyses (Panel A: the episodic future thinking task (EFT), the episodic memory task (EM), the scene construction task (SC), and the combination of all three tasks (ALL)); for the CEN and ‘Spatial Coherence Index’ (SCI) analyses (Panel B: EFT, EM, SC and ALL); for the CEN and ‘Vividness’ analyses (Panel C: EFT, EM, SC and ALL); for the CEN and ‘Sense of Presence’ (SoP) analyses (Panel D: EFT, EM, SC and ALL); and for the CEN and ‘Detail’ analyses (Panel E: EFT, EM, SC and ALL).

Figure 5 also highlights the lack of consistency observed across our two cohorts with respect to the relationship between the EI and CEN. The EI score is a composite comprised of a number of individual parameters, some of which were scored by the experimenter and others that were self-rated by the participants. In order, in part, to consider whether a lack of blinding may have influenced Cohort 1 data, we further explored the influence of CEN on these individual parameters, particularly those not readily influenced by scorer bias i.e. participants’ ratings of their subjective experience recollecting/imagining the specific events/scenarios (see Table 2 for more details). We asked whether consistent correlations between CEN and any of these individual self-rated parameters would be evident across our two cohorts. Of specific interest, were the questions vividness (i.e. “How vivid was the event you recollected/imagined in your mind’s eye?”) and sense of presence (SoP; i.e. “How much of a sense of being there did you have when recollecting/imagining….etc?”), as both of these questions feed directly into the EI score. For completeness, the three remaining questions (i.e. difficultly, level of detail and emotional valence) were also explored.

Significant negative correlations were evident between CEN and participants’ vividness ratings, in both Cohort 1 and Cohort 2, for imagined events (i.e. EFT and SC trials), and for the ‘EFT+EM+SC’ composite scores. There was consistent lack of evidence of a relationship between CEN and the vividness of participants reported episodic memories (see Table 4). Hence, higher CEN consistently related to less vivid imagined, but not recollected, scenarios.

A similar pattern of negative correlations between CEN and participants’ sense of presence in their simulated future thoughts (EFT) and scenes (SC) in both cohorts with the exception of Cohort 2 EFT SoP (p = 0.061; see Table 4); whereby higher CEN consistently related to an attenuated sense of personal presence within imagined, but not recollected, scenarios.

Self-reported perceived level of detail within the imagined showed less consistency across cohorts, whilst perceived task *difficulty* only correlated with CEN on Scene Construction trials for Cohort 1.

Finally, higher CEN correlated significantly with lower emotional valence of imagined (i.e. EFT and SC trials; EM trials p = 0.051). Unfortunately, as this latter parameter was only assessed in Cohort 2, it is not possible to assess the between-cohort consistency (for full details see Table 4).

Hence, consistency across cohorts was evident for many of these individual markers of participants’ subjective experience imagining future thoughts or atemporal scenes; most notably for the perceived vividness and sense of presence that participants reported experiencing when imagining future or fictitious scenarios. There was also notably consistent null results for recollected events (with the possible exception of Vividness and Emotional Valence in episodic memories for Cohort 2). This division between imagined and recollected events was not anticipated a priori. Importantly, this division was reinforced when the data for both cohorts was analysed as a whole (i.e. Cohort 1+2) (see Table 4).

As the above dichotomous statistical outcomes suggest a division between the memory and imagination domains, we explored visual representations of each point and interval estimate for the three parameters that showed the most consistent relationships with CEN i.e., vividness, SoP and level of detail (see Figure 5: Panels C-E). This illustrated a number of potentially relevant points of interest. Firstly, and almost without exception, the point estimates across all tasks and cohorts consistently fell below zero. Moreover, the interval estimates in Cohort 1 were substantially wider than in Cohort 2 for all three relationships, indicating an increase of precision in the larger of the two samples (i.e. in Cohort 2). However, interval estimates within the Cohort 1 analyses were widest for the EM estimates, indicating less precision in these estimates. This reduced precision may have reduced the likelihood of observing a significant correlation between CEN and the individual parameters in the EM task in the combined cohort analysis – an observation that may be particularly relevant for the vividness parameter (as here the point and interval estimates are highly consistent across tasks in Cohort 2; Figure 5C). The influence of this reduced precision in the EM estimates in Cohort 1 on the combined cohort analysis is less clear for the SoP (Figure 5D) and level of detail (Figure 5E) analyses where a decrease in precision does not appear to account for the lack of a significant correlation between CEN and these parameters for the EM (but not the EFT and SC) task.

Finally, unlike CEN, BDI-II scores showed relatively few significant correlations with individual dependent variables. In fact, no significant correlations were observed between BDI-II scores and any parameters in Cohort 2 (see Table 4). However, in Cohort 1, BDI-II scores correlated with EI for SC and the composite (EFT+EM+SC) measure, with vividness and detail for the composite measure, and with SoP in EFT, SC and the composite score. When the two cohorts were combined, BDI-II scores correlated with SoP for SC only (see Table 4).

#### Mediation Analyses (Phase 2)

The SPSS macro PROCESS v3.5 (Hayes, 2018) was used to perform simple mediation analyses to assess the extent to which BDI-II scores mediated the relationship between CEN and the relevant dependent variable. Given our hypotheses with respect to depression, BDI-II scores were used as a mediator.

##### Cohort 1 Data

Mediation analyses were conducted to assess whether BDI-II scores mediated the relationships between CEN and the dependent variables that were found to correlate with both CEN and BDI-II scores. In all analyses, it was concluded that no mediation occurred (See Supplementary Material Part 6 for details).

##### Cohort 2 Data

As there were no instances where BDI-II scores correlation with the dependent variables in the Cohort 2 dataset, no mediation analyses were performed.

##### Combined Data (Cohorts 1 and 2)

When both CEN and BDI-II scores correlated with an individual parameter/dependent variable simple mediation analyses were performed. When the date from both cohorts was analysis collectively, there was only one instance in which both CEN and BDI-II scores correlated with an individual parameter/dependent variable - SC SoP scores (see Table 4). For completeness, simple mediation analyses were performed even when BDI-II correlations were only significant at the p < 0.1 level. No significant indirect effects of BDI-II scores on the relationship of CEN with the dependent variables were observed (see Figure 6).

**Figure 6:**
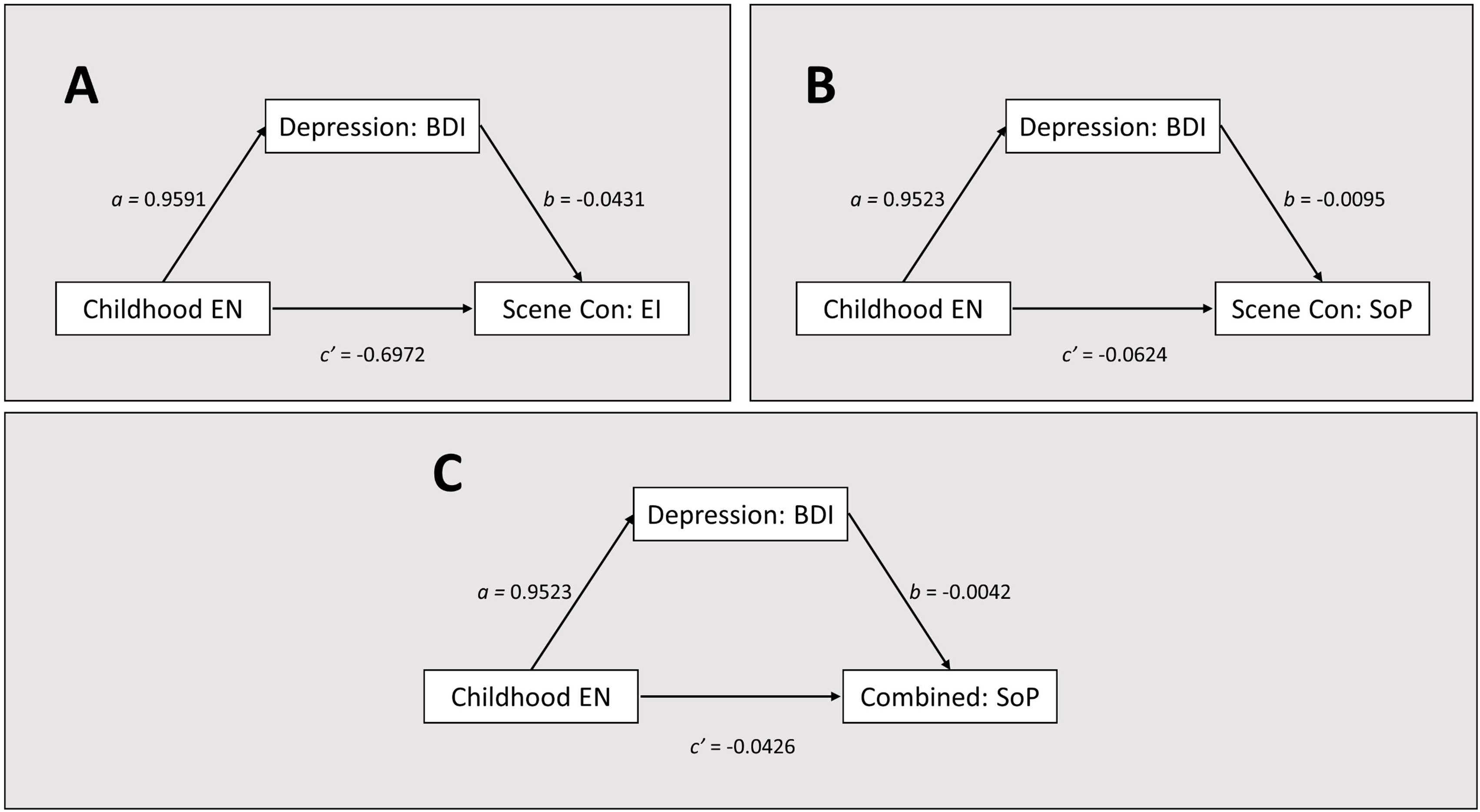
Phase 2 Mediation Analyses (Cohorts 1 + 2). <u>Panel A</u>: Mediation analysis concluded that BDI scores did not mediate the relationship between CEN and Scene Construction Experiential Index when the data from both Cohorts 1 and 2 were combined. This was also observed when the Cohort 1 and Cohort 2 datasets were analysed separately (Cohort 1, Indirect effect: B=-.2522, CI [-.7474, .1420]; Cohort 2, Indirect effect: B=.0254, CI [-.2205, .3669]). <u>Panel B</u>: Mediation analysis concluded that BDI scores did not mediate the relationship between CEN and Scene Construction Sense of Presence when the data from both Cohorts 1 and 2 were combined. This was also observed when the Cohort 1 and Cohort 2 datasets were analysed separately (C1 Indirect effect: B=-.0312, CI [-.0734, .0022]; C2 Indirect effect: B=-.0057, CI [-.0367, .0157]). <u>Panel C</u>: Mediation analysis concluded that BDI scores did not mediate the relationship between CEN and Sense of Presence scores (combined for EFT, EC and EM) when the data from both Cohorts 1 and 2 were combined. This was also observed when the Cohort 1 and Cohort 2 datasets were analysed separately (C1 Indirect effect: B=-.0242, CI [-.0671, .0071]; C2 Indirect effect: B=.0021, CI [-.0284, .0147]).

### Phase 2 (Cohort 2 only): Performance on laboratory-based tasks of anterograde memory and IQ

The additional cognitive tasks included in Phase 2 for Cohort 2 participants assessed predicted IQ, verbal memory (stories recall), visual-spatial memory (ROCF recall), and the paired-associates memory. CEN did not significantly correlate with any of these measures apart from the ROCF visual-spatial memory recall (see Table 5). In this task, participants were required to copy the complex figure and to reproduce this figure from memory approximately 30 minutes later. Accuracy of the delayed recall figure was then compared to the participants’ initial copy accuracy. CEN significantly negatively correlated with copy accuracy (r_s_=-0.413, [-0.691, -0.072]), delayed recall raw score (r_s_=-0.529, [-0.741, -0.242]), delayed recall/copy accuracy (r_s_=-0.475, [-0.707, -0.160]). Hence, participants who reported higher levels of CEN tended to be worse at both copying and recalling the figure. This latter deficit was evident even when recall scores were adjusted to account for poorer copy accuracy. As BDI did not correlate with any of the above variables, no mediation analyses were performed.

**Table 5.** *(Cohort 2 only)*: Relationships between CEN, Wellbeing, and neuropsychological assessments of anterograde memory and cognitive ability in Cohort 2 participants (n=41): Phase 2. *p<.05, **p<.01, ***p<.001. Shaded findings also indicate significant results. ROCF = the Rey-Osterrieth Complex Figure task, Story A/B = Logical Stories subtest (Wechsler, 1997), PA = Paired Associates (see methods), WTAR = Weschler Test of Adult Reading.

|  | CEN |  |  | BDI-II |  |  |
| --- | --- | --- | --- | --- | --- | --- |
| | $r_s$ | Sig. | 95% CI | $r_s$ | Sig. | 95% CI |
| ROCF Copy | -.394 | .011* | -.66, -.05 | .022 | .893 | -.37, .38 |
| ROCF Delayed | -.536 | .000*** | -.74, -.23 | -.177 | .274 | -.53, .19 |
| ROCF Accuracy | -.486 | .001** | -.71, -.17 | -.165 | .310 | -.52, .19 |
| ROCF Recognition | -.047 | .775 | -.39, .31 | .170 | .300 | -.13, .47 |
| Story A Immediate | -.155 | .335 | -.46, .19 | -.171 | .293 | -.50, .16 |
| Story B Immediate 1 | -.196 | .220 | -.48, .13 | -.057 | .726 | -.41, .29 |
| Story B Immediate 2 | -.149 | .353 | -.44, .19 | -.220 | .173 | -.53, .17 |
| Story A Delayed | .074 | .647 | -.25, .36 | -.119 | .466 | -.44, .22 |
| Story B Delayed | -.053 | .743 | -.38, .26 | -.018 | .914 | -.36, .39 |
| Paired Associates T1 | -.125 | .444 | -.46, .21 | -.016 | .921 | -.36, .32 |
| PA Criterion | -.121 | .457 | -.22, .41 | -.108 | .512 | -.45, .25 |
| PA Delayed | -.165 | .309 | -.46, .17 | -.158 | .337 | -.48, .20 |
| WTAR-IQ | .051 | .751 | -.26, .36 | -.295 | .065 | -.55, .01 |

## Discussion

We conducted two independent (Cohort 1 and Cohort 2), biphasic (Phase 1 and Phase 2) studies using non-clinical samples of young-adult undergraduate university students. Phase 1 was conducted online. Here participants completed the CTQ-EN subscale and answered questions pertaining to their current wellbeing. CTQ-EN scores were then used to generate subsets of participants whose self-reported levels of childhood emotional neglect (CEN) straddled the full range of this subscale. Phase 2 was conducted in the laboratory. Here these subsets of participants performed a neuropsychological/cognitive task battery.

Across both phases, and in each cohort, we sought to explore the relationship between retrospectively self-reported levels of CEN and current wellbeing/depression severity. In keeping with the well-documented relationship between higher levels of CEN and poorer current wellbeing in clinical populations, we predicted a negative correlation between these two parameters. In addition, we sought to elucidate the relationships between both CEN and current wellbeing/depression scores with respect to performance on tasks that tap into key hippocampal-related cognitions such as episodic memory recollection, episodic future thinking and scene construction (Cohorts 1 and 2), and performance on standardised neuropsychological assessment of anterograde memory (Cohort 2). In both cohorts, we sought to determine whether task performance related to levels of self-reported CEN, or to current wellbeing/depression, or to both. Where evidence of the latter was present, we used mediation analyses to establish whether current wellbeing/depression scores mediated the relationship between CEN and the parameter in question. In this way we sought to garner evidence, at a cognitive level, to inform the current debate regarding the antecedents of the neurobiological (e.g. reduced hippocampal volume) and cognitive (e.g. increased over-general memories/future thoughts) differences observed in psychiatric disorders such as MDD and PTSD, and in individuals who have experienced emotional neglect in childhood. We predicted that if these neurobiological and cognitive differences are initially triggered/driven by the childhood adversity, but later compounded by disorders such as MDD, then task performance would relate to both CEN and current depression scores.

### Childhood emotional neglect and current wellbeing in young adult University students

As hypothesised, higher levels of self-reported CEN were consistently related to poorer current wellbeing. In addition, higher levels of CEN were also related to higher scores on the BDI-II. Hence, young adult university students who retrospectively self-reported higher levels of childhood emotional neglect also tended to rate their current wellbeing as poorer and score higher on a depression severity scale than their peers who reported lower levels of CEN. Higher levels of state anxiety also related to higher levels of CEN. Given the extensive previous research showing a clear link between both prospective and retrospective reports of childhood maltreatment and increased incidences of subsequent psychopathology, this is unsurprising. It does however, extend this observation into multiple samples of high-functioning young-adult i.e. current university students, who were recruited to the study on the basis of their university registration, as opposed to their clinical psychiatric history (as is more typical in previous studies). It is also important to document that even in this high-functioning, young adult sample, which likely contain individuals who have yet to experience mental health difficulties and/or those whose mental health difficulties are emerging, this relationship is still evident and robust.

### Childhood emotional neglect and hippocampal-dependent cognitive function

As hypothesised, higher levels of self-reported CEN were also associated with poorer in-person (Phase 2) task performance on a range of measures designed to assess hippocampal-dependent cognitive processes such as recollecting personally experienced events, simulating ones’ personal future, and imagining atemporal scenes. More specifically, in Cohort 1, higher levels of CEN were related to reductions in the overall richness (i.e. EI scores) of simulated future scenarios (EFT), recollected personal past experiences (EM), and imagined atemporal fictitious scenes (SC). We did not observe the anticipated correlation between CEN and the spatial coherence of imagined future or fictitious atemporal events (i.e. scenes) or recollected memories in this cohort. This null result was also observed in Cohort 2, and when the data from both cohorts was combined.

We did not replicate these strong correlations between CEN and EI in Cohort 2, perhaps due to a lack of blinding of experimenters/raters in Cohort 1 (a potential confound that was corrected in Cohort 2). However, consistency was observed across cohorts for many of the parameters characterised by participants. More specifically, in Cohort 1 higher levels of CEN was associated with reduced vividness and level of detail, and a diminished sense of presence (SoP) when participants imagined future events (EFT) and constructed fictitious atemporal scenes (SC). These correlations were not observed when participants recollected past events in which they were personally present (EM; see Table 4). In Cohort 1, higher levels of CEN also related to increased difficulty constructing fictitious scenes. In Cohort 2, higher levels of CEN related to lower vividness (EFT, SC, and ‘EFT+EM+SC’), attenuated SoP (SC and ‘EFT+EM+SC’), and reduced emotional valence (EFT, SC, and ‘EFT+EM+SC’). Hence, across two independent cohorts, we observed evidence that individuals who self-report higher levels of CEN also report mental simulations that are less vivid and in which they feel less personally present. This effect appeared to be more pronounced, and more consistently observed, in the imagination (and specifically in constructed scenes) as opposed to the memory domain.

Given the extensive literature relating both to early life chronic stress and psychopathologies such as MDD to over-general memories/future thinking, it is perhaps unsurprisingly that CEN ratings, in addition to correlating with estimates of current wellbeing, also consistently related to parameters such as vividness and sense of presence. Notably also was the relationship between higher levels of CEN and higher levels of emotional detachment from simulated future events and scenes (although this parameter was only assessed in Cohort 2), which resonates with the core features of over-general memory/future thinking, i.e. an attenuation of specificity and a general detachment within descriptions of episodic events.

### Current wellbeing/depressive symptomology and hippocampal-dependent cognitive function

Contrary to our hypothesis (and previous findings), fewer significant correlations were observed between BDI scores and these memory and imagination parameters, and no correlations were observed with these parameters and estimates of current wellbeing. With respect to BDI correlation in Cohort 1, lower BDI scores related to reduced SoP for EFT and SC trials (and for the combined ‘EFT+EM+SC’ scores) but not for EM recollection. Lower BDI scores were also related to attenuated overall scene richness (i.e. EI scores) but only for SC trials (and ‘EFT+EM+SC’) and only within the Cohort 1 dataset . In addition, it was only when the data for the EFT, SC and EM trials were combined that more consistent correlations were observed (BDI and EI, vividness, detail, and SoP were found; see Table 4), however this was not evident in the Cohort 2 data where no correlations with any parameter and BDI scores were reported in Cohort 2 (including for the emotional valence ratings assessed here; see Table 4).

### CEN, Current wellbeing/depressive symptomology and hippocampal-dependent cognitive function

Given the established relationship between depression and phenomena such as reduced hippocampal volume and OGM, and the robust relationship between higher levels of CEN and poor current wellbeing that we consistently observed here (and is well documented in the literature), these relationships warranted further investigation. We thus conducted mediation analyses in all instances where a specific parameter was found to correlate with both CEN and current depression (i.e. BDI-II scores; see Table 4). We asked whether CEN had an effect the specific parameter via current depression. No significant effects of the mediation pathways were found, indicating that the relationships between CEN and each dependant variable (e.g. SoP (SC)) and between current depression scores and the same dependant variable (e.g. SoP (SC)), occurred independently of one another. Whilst caution is needed when interpreting any null result, especially with relatively low numbers of participants, future work should endeavour to replicate these findings with larger participant numbers, and to unpack the relationship between these variables further. Notably however, combining the data across the two cohorts did not alter results (see Table 4, Cohorts 1+2) indicating that our current findings do not support the hypothesis that mediation via current depression best explains the association between higher levels of CEN and the attenuation of subjective experience of simulated mental events.

### Imagination versus Recollection

It is not immediately clear why, in both cohorts, we observed greater consistency with respect to the relationship between CEN and the vividness and SoP with participants’ imagined scenes and imagined personal futures (EFT), as opposed to the qualia of their episodic memory recollections (EM). This dissociation between the imagination and the recollection domains was not hypothesised a priori. In Cohort 2, more traditional memory measures that are reliably impaired in individuals who suffer selective, extensive and bilateral hippocampal damage and amnesia were included in Phase 2. Whilst we observed a relationship between CEN and impaired complex figure recall accuracy (RCOF), we did not observe a similar relationship between CEN and performance on the other memory tasks (e.g., story recall and verbal paired associate learning). This pattern of findings is consistent with the suggestion that the impact of CEN is more pervasive in the imagination domain, and resonates with data in depression showing more severe deficits in EFT than EM (Addis et al., 2016).

Moreover, the hypothesised correlation between CEN and the spatial coherence (SCI) of the recollected/simulated events was not evident in either dataset, nor was it evident when the data from both cohorts was combined, indicating that the spatial integrity of the participants’ simulated/recollected scenarios did not relate to self-report levels of CEN (or BDI scores). Scene Construction Theory posits that the ability to simulate spatially coherent scenes affords us the ability to re/pre-play past and future scenarios within a spatially realistic backdrop. If CEN affects the vividness, detail, and sense of presence within simulated scenes, but does not necessarily impact on the spatially contiguousness of these scenes, then this may explain why attenuated performance on some (e.g., the ROCF task), but not all (e.g., verbal story recall and verbal paired associates) anterograde memory tasks related to CEN. In other words, whilst these scenes may be degraded in terms of their vividness, detail…etc., their basic structural backdrop appears to be preserved. This basic structure may be sufficient to support mnemonic performance on tasks such as story recall and paired associates, as encoding and recollecting such material can likely be achieved without the vivid reconstructions of real-world spaces. Why this basic structure appears to be more robustly able to support the replay of personally experienced events, than the pre-play of simulated personal future events (i.e., EFT), is unclear. Notably here, it was on the scene construction trials in which we found the most consistent relationship between CEN and Vividness/SoP, suggesting that scenarios in which participants are explicitly required to construct without reference to lived experiences, are those that appear to be most problematic for participants with higher levels of CEN. Future studies exploring the possibility that specific deficits in the imagination of spatially coherent scenes (in addition to deficits in parameters such as vividness and SoP) are needed. Moreover, further studies aimed at deconstructing, and more thoroughly exploring, the phenomenological experience of episodic memory recollection and episodic future thinking in relation to CEN are required to help establish whether subtle differences in the domain of memory do exist but were not adequately quantified by the methodology employed here; specifically with respect to how such events are re-lived and subjectively experienced. Such studies could also explore the influence of event valence on these parameters. Additionally, the unpredicted association between ROCF copy accuracy and CEN may indicate possible difficulties in attentional or visuospatial processing in those with CEN histories.

As with the unexpected dissociation between the memory and imagination domains, we did not anticipate observing differences in parameters such as simulation scene richness (EI)/vividness/level of detail/SoP/emotional salience, whilst simultaneously observing no differences with respect to how participants reported the spatial coherence of these simulations. This contrasts with previous neuropsychological findings that have consistently shown a tight coupling between these features. More specifically, and relative to healthy controls, patients with bilateral hippocampal damage (Hassabis et al., 2007; Mullally et al., 2012), bilateral posterior parietal cortex lesions (Berryhill et al., 2010), unilateral prefrontal cortex (PFC) lesions (Berryhill et al., 2010), Alzheimer’s Disease (Irish et al., 2015), and Posterior Cortical Atrophy (Ramanan et al., 2018) all show severe impairments in *both* overall scene richness i.e. EI (a measures that includes parameters such as the vividness of the simulated experience), and the spatial coherence (i.e. SCI) of these (re)-constructed events. Moreover, and to the best of the authors’ knowledge, deficits in scene construction (e.g. EI, Vividness, SoP, Level of Detail) in a context of preserved SCI have not been observed in any neurological population studied (although see Berryhill et al., 2010, for discussion of a potential dissociation between EI and SCI deficits evident only when the SC performance in patients with bilateral posterior parietal cortex damage is directly compared with SC performance in patients with bilateral hippocampal damage, as opposed to with healthy controls). It is therefore challenging to localise which component(s) of the scene construction/episodic future-thinking network are specifically impacted by high levels of childhood emotional neglect, particularly given that this unexpected dissociation between these phenomenological features and the spatial cohesiveness and integration (SCI) of the imagined scenario has not previously been reported in a neuropsychological studies. Future studies should seek to explore the neural correlates these dissociations, as this could inform current thinking on the neural mechanism underpinning the emergence of mood disorders within the context childhood adversity.

### Online versus laboratory-acquired data

The results of the additional online component conducted in Cohort 2 (Phase 1), at least on initial inspection, appear to be more consistent with the original hippocampus-orientated hypothesis. More specifically, participants in this cohort (n=456) reported higher levels of CEN, also reported having lower confidence in their ability to remember specific past events and greater difficult remembering learnt information (alongside greater difficulty imagining future events and how they might feel in these future scenarios, and greater difficulty visualising images/pictures in their mind’s eye). Given the relative paucity of consistent effects in the domain of memory observed in the Phase 2 data, these significant correlations are noteworthy. We also reported that current wellbeing appeared to mediate the relationship between CEN and each of these parameters, with the exception of CEN and ease of mental imagery. This pattern of results is notably distinct from both Phase 2 Cohort 1 and Phase 2 Cohort 2 data, which consistently failed to find a significant indirect effect of the mediator variable (BDI scores) on the relationship between CEN and the respective parameter.

One possible explanation is that the increased power associated with the larger numbers of participants that it is possible to test online, versus in the laboratory, may indicate that the observed relationships in our laboratory findings were spurious in nature (and the absence of a mediation effect driven by a lack of power). However, it is also possible that participants were less accurate when rating their abilities online, as in these instances participants would likely have been doing so without an objective example event salient in their mind. This differs from participants in the laboratory-based phase of both cohorts, whereby ratings were acquired on a trial-by-trial basis; immediately after simulating/recollection an event. Similar differences between objective and subjective scores have been noted in the navigation literature. For instance, when asked to rate their navigational abilities prior to an experiment commencing, participants self-ratings had little relationship with their subsequent navigational performance during the experiment (Heth et al., 2002), but that when the ratings were acquired after the task had been performed, the ratings did reflect performance. Another important difference between the results of the online and laboratory-acquired data, is that poorer wellbeing ratings were associated with all variables rated online i.e. with lower subjective ratings of memory (both episodic and semantic), poorer future thinking abilities, and increased difficulty visualising mental images/pictures. This was not observed in the laboratory-based (Phase 2) datasets, where no correlations with current wellbeing and relatively few significant correlations with BDI scores (see Table 4). One possible interpretation of this difference is that more generalised subjective-ratings of abilities may be more biased by current wellbeing, than ratings acquired using a trial-by-trial task-based method, leading to a more generalised depression of all ratings scores in participants with poorer wellbeing.

Hence, caution may be required when directly comparing our online ratings (that were not related to a specific instance of recollection/simulation) and ratings acquired immediately after actual task performance in a laboratory setting as it is likely inappropriate to consider them as equivalent measures of the same underlying process. Instead, both the online and laboratory-based findings offer differing insights into how CEN and current wellbeing related to our subjective experience of memory/imagination/future simulation at both a general level and at a more task-specific level. The fact that current wellbeing appears to play a more influential role in the generalised self-report ratings is perhaps unsurprising given the wealth of clinical and research evidence, relating patterns of low self-esteem and negative self-constructions with vulnerability for depression (e.g. Orth et al., 2009; Sowislo & Orth, 2013).

### Overarching findings

There are, however, some interesting parallels between the specific parameters that appear to relate to CEN identified in the extended online component (Phase 1, Cohort 2) and the laboratory components (Phase 2, Cohort 1 and 2). More specifically, when these are considered together, higher-levels of CEN were most consistently related to parameters that specifically tapped into the subjective/phenomenological experience of episodic imagination i.e., such as vividness and sense of presence. These parameters may relate to the level of emotional presence the participants experienced during these simulations. Less consistent correlations were observed with respect to the subjective experience of memory, whereby significant correlations were evident in Phase 1, Cohort 2 (e.g. confidence in recollection ability) but not significantly in the Phase 2 in-person data. CEN also did not consistently correlate with more objective measures of anterograde memory (e.g., number of word pairs or story details correctly recollected).

This constellation of effects shares similarities to that observed in patients following bilateral parietal lobe lesions (Berryhill et al., 2007; Berryhill et al., 2010; Simons et al., 2010); a profile that is distinct from that observed following hippocampal/medial temporal lobe damage. More specifically, patients with bilateral hippocampal damage are typically not considered to be amnesic (e.g. Ally et al., 2008; Simons & Mayes, 2008; Simons et al., 2008) but are impaired (e.g., they provide a lack of detail) when asked to freely recall autobiographical memories (Berryhill et al., 2007; see also Davidson et al., 2008). The possible neural underpinnings of this unexpectedly circumscribed pattern of findings, and the potential implications for psychopathologies driven by childhood adversity, is explored in further detail elsewhere (Watson and Mullally, Paper B). Future neuroimaging studies will also assist in elucidating any specific role played by this region, both in response to the experience of CEN, and as a potential precursor to the development of future psychopathology. However, whilst assessing brain structure and function is clearly important, the importance of also assessing higher-order cognitive functions, such as the parameters measured above, should not be overlooked. We argue that the results of the cognitive studies reported here may provide important guidance when attempting to disentangle both neurocognitive consequences of aversive childhood experiences such as CEN (for further discussion see Watson and Mullally, Paper B).

### The qualia of episodic simulation as a potential therapeutic interventions

Given the well-established significance of CA, including emotional neglect, in shaping population level mental health (McLaughlin et al., 2014), developing preventative therapeutic interventions following the experience of CA should be a priority. Given the wide range of cognitive processes impacted by CA, these will likely be varied and multifaceted. However, the experiential differences in the qualia of episodic simulations linked here to the experience of CEN may represent an important therapeutic target to decrease CEN-driven latent vulnerability to psychopathology.

Interestingly, a small number of intervention studies have been conducted which have attempted to modify OGM in patients who have existing depression using a group-based cognitive intervention, MEmory Specificity Training (MEST) (Neshat-Doost et al., 2013; Raes et al., 2009). Raes et al. (2009) Raes et al. (2009) found that after four consecutive weekly 1-hour MEST sessions, the retrieval style of patients [who were currently receiving inpatient treatment for depression (n=10; all female)] became significantly more specific (and hence less over-general). Whilst improvements in memory specificity did not appear to relate to changes in depression, no follow-up was conducted. This was addressed by Neshat-Doost et al. (2013) who performed a blinded, randomised design conducted in depressed Afghan adolescents who had all lost their fathers due to the war in Afghanistan and had subsequently immigrated to Iran as refugees (n=12 MEST group, n=11 control group; mean age = 14.88; 11 girls and 12 boys). Following 5-week MEST program, those who received MEST showed a significant change in the proportion of specific memories they generated. This change in memory specificity also predicted follow-up (2-months later) depressive symptomology, over and above baseline depression, and mediated the relationship between participating in MEST and the reduction in subsequent depressive symptoms.

Hence, if reducing OGM appears able to drive subsequent improvements in adolescents already experiencing symptoms of depression, it is plausible that targeting OGM and over-general episodic cognition in children and/or adolescents who have experienced CEN, but who have not yet developed a mood disorder, may help to assuage subsequent vulnerability. This is supported by findings linking the ability to generate specific future simulations with greater psychological wellbeing, problem solving and coping behaviours (Addis et al., 2016). In addition, the findings reported here may also have implications for the use of imagery-based therapies for depression (e.g. Blackwell & Holmes, 2017); particularly when considering whether imagery-based therapies are appropriate for those with CEN histories, and/or whether imagery-based therapies may need to first include a mental imagery/scene-construction-training component for individuals who cannot readily conjure up vivid mental images.

### Retrospective versus prospective measures of childhood adversity

Finally, there remains an important debate with regard to whether retrospective or prospective measures of childhood adversity are more appropriate to use in such studies. Retrospective self-reports of childhood maltreatment have been queried for unreliability and recall bias; in particular, mood-congruency effects. However, by fitting a structural equation model to repeated measures of retrospective self-reports of childhood maltreatment and current mental health (when participants were aged 18 and repeated at age 21), Fergusson and colleagues found that recall bias accounted for less than 1% of reported variance, and that overall errors of measurement in childhood maltreatment did not threaten study validity (Fergusson et al., 2011).

Prospective measures of CEN, and childhood adversity more generally, do offer obvious benefits to longitudinal studies attempting to decouple temporal and causal relationships between childhood adversities and the onset of psychopathologies. For instance, they enable the objective quantification of CEN/adversity by an outsider (e.g., by a social worker…etc.). However, retrospective measures, such as those utilised here, also offer important advantages (Baldwin et al., 2019). For instance, a strength of the EN subscale of the CTQ is that although it is a subjective, retrospectively-viewed measure, which is influenced by current social circumstance and emotional wellbeing (and all the complexity that this affords it; Colman et al., 2016), it also enumerates an individual’s current perception of the care that they received in childhood. This perception may, in itself, capture something important and different to objective ‘other’ generated ratings. Interestingly, whilst both prospective informant-reports (obtained from caregivers, researchers and clinicians) and retrospective self-reports of childhood maltreatment (obtained using the CTQ) were predictive of a range of psychiatric problems at age 18, the strongest associations were observed for retrospective self-reported childhood maltreatment (Newbury et al., 2018). Hence, retrospectively self-reported childhood maltreatment may measure a construct that taps into an individual’s own current perspective/recollection of their childhood experiences; and it may be this perception that captures something that is particularly relevant to psychopathology.

In addition, the pattern of results in Phase 2 is not consistent with the counter-argument that current emotional wellbeing/mood systematically biased participants’ self-report retrospective CEN ratings, as if that was the case, then the pattern of correlations observed for CEN and current wellbeing/BDI scores should mirror one another. This was not the case, nor did we find any evidence of mediation (via BDI scores) in the laboratory-acquired data. Hence, it appeared that CEN estimates captured something unique and not adequately captured by the wellbeing questions/BDI questionnaire utilised here. Whether this is a veridical account of childhood experiences, or some important additional marker of current wellbeing, is an open, and interesting, question.

## Conclusion

We have examined two cohorts of university students and demonstrated a consistent correlation between retrospective self-reports of childhood emotional neglect (CEN) and current wellbeing, and between CEN and depressive symptomology, in two non-clinical young-adult samples. These relationships were apparent in both the large online data collection phase (Cohorts 1 and 2 - Phase 1), and in the smaller laboratory gathered data (Cohorts 1 and 2 - Phase 2). Moreover, in these later in-person datasets, we demonstrated consistent correlations between higher levels of CEN and the reduced vividness and sense of presence when participants simulated fictitious and/or future based events in which they were personally present, but not consistently when participants recollected personal past events. No relationship between CEN and the reported spatial coherence of participants’ simulations or recollections were observed, which in combination with the absence of a strong effect in the domain, argue against a selective hippocampal-driven account of CEN. The results of mediation analyses found no evidence to support the hypothesis that mediation via current depression accounted for the observed relationship between CEN and impoverished components of simulated events. In addition, higher levels of reported CEN was associated with poorer performance on visuospatial, but not verbal, tasks of anterograde memory (Cohort 2 - Phase 2).

Some differences were observed in the relationships between CEN, current wellbeing, and subjectively-reported estimates of hippocampal functions when data was gathered online (Cohort 2 – Phase 1) as opposed to in the laboratory; with such measures appearing to relate more consistently correlations with current wellbeing than laboratory-based measures. Despite this, higher estimates of CEN were still found to be related to poorer self-reported mental imagery abilities, and we found no evidence that mediation via current wellbeing accounted for this relationship.

### Context

In multiple non-clinical samples of high functioning young adults (currently attending university), we consistently observed a link between CEN and variables that tap into the vividness of mental simulations. We conclude that the putative role of this specific constellation of impairment may play an important role in mediating the risk of depression in individuals who have experienced high levels of childhood emotional neglect. As such, future studies, further characterising these cognitive differences are necessary to enable a more nuanced understanding to the specific consequences of childhood maltreatment on the developing brain, and how and why, this confers an increased risk of developing a subsequent psychopathology.

## Data Availability

All data produced in the present study are available upon reasonable request to the authors.

