## Supplemental Materials for "Growing up unloved: the enduring consequence of childhood emotional neglect on the qualia of memory and imagination"

**Supplementary Material – Part 1**

### Histograms of CEN scores for Cohorts 1 and 2

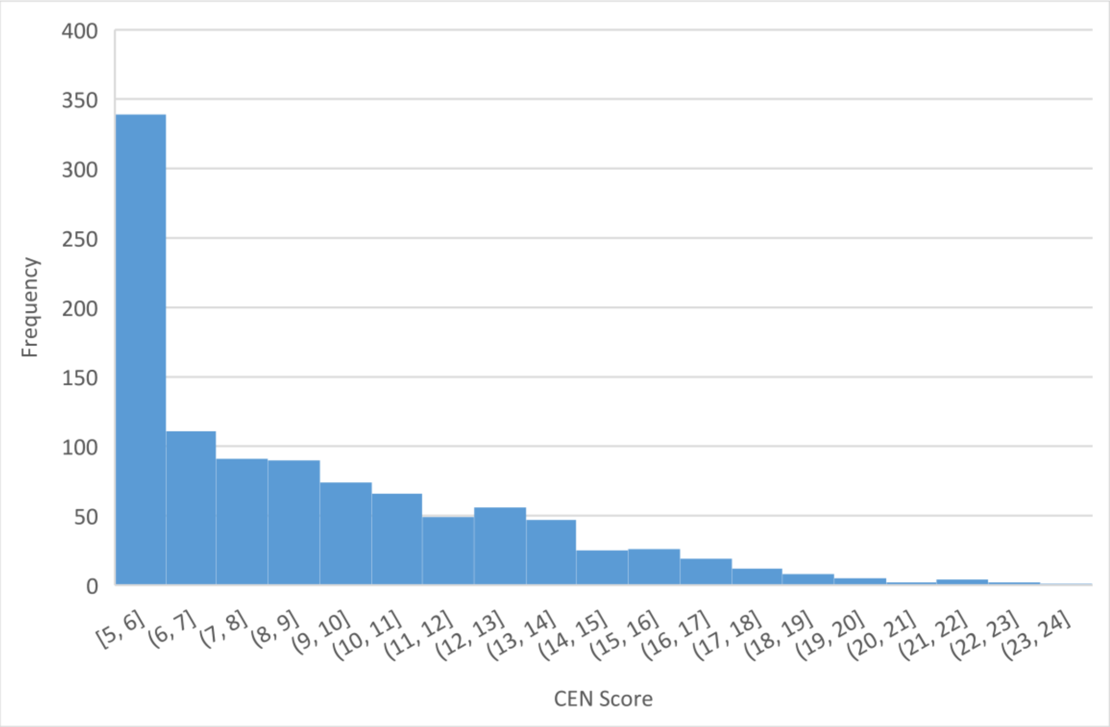


**Figure S1**. Cohort 1: Histogram of participants' CEN scores. Maximum possible range of scores = 5-25.

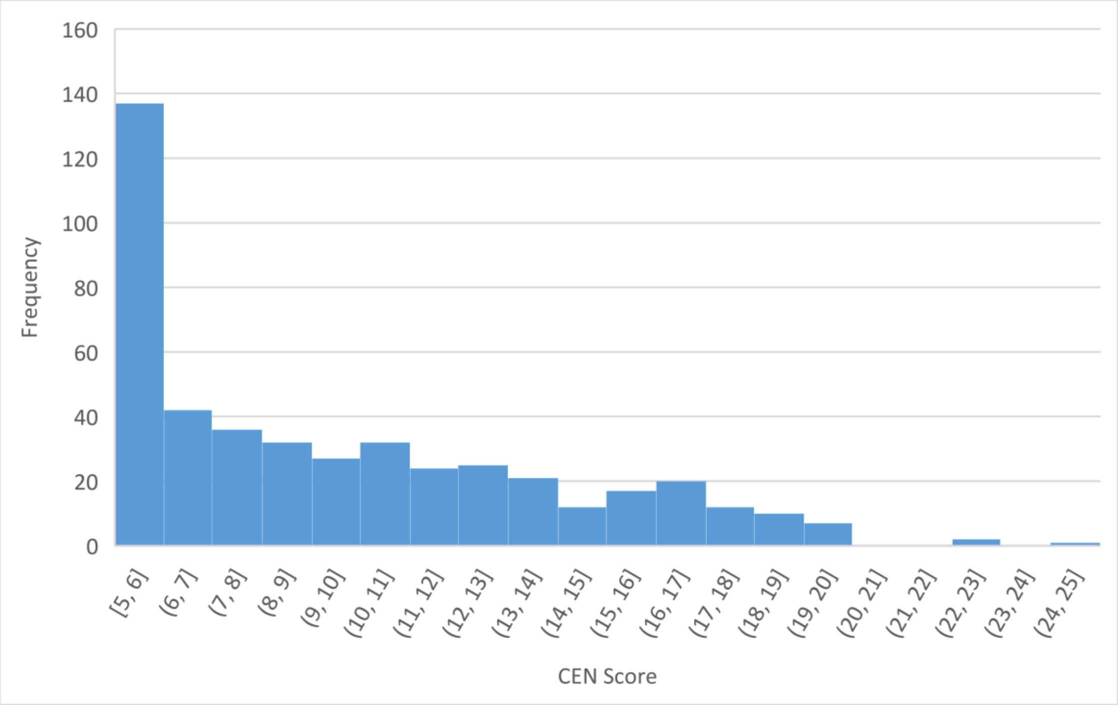


**Figure S2**. Cohort 2: Histogram of participants' CEN scores. Maximum possible range of scores = 5-25.

**Supplementary Material – Part 2**

### Anterograde Memory Tasks Procedures

#### *The Rey Osterrieth Complex Figure (Osterrieth, 1944)*

Participants were initially required to copy the complex figure, composed of 18 geometric shapes, in as much detail as possible. After thirty minutes they were instructed to re-draw the figure from memory (Delayed Recall). To prevent active rehearsal of the figure during the delay period, participants were not informed that there would be a recall component to this task prior to receiving the instruction to re-draw the figure from memory. Both the copy and delayed recall drawings were scored (0-2/unit; maximum score=36), and an overall accuracy score (%) was calculated (Copy/Delayed *100).

#### *Logical Memory (WMS-III, Weschler, 1997).*

This task consists of two short stories (Story A and Story B), each comprised of 25 verbal units. Story A was read aloud by the examiner and immediately recounted by the participant in as much detail as possible [Immediate Recall: Story A]. This process was repeated for Story B [Immediate Recall: Story B(1)], followed by a second reading and recollection cycle of Story B [Immediate Recall: Story B(2)]. After a thirty-minute delay, participants are again asked to recollect both stories in as much detail as possible [Delayed Recall: Story A, Story B]. As with the Rey Osterrieth Complex Figure task, participants were not informed of the delayed recall phase prior to receiving the instruction to recollect each story at delay. Both stories were scored using the WMS-III scoring criteria whereby a single point was awarded for each unit correctly recollected.

***Paired Associate Task***

Participants were presented with 24 unrelated word pairs (e.g. BANNER MONK) displayed centrally on a computer screen for four seconds/word pair. Participants were instructed that at later time they would be presented with the left-hand word of the pair and asked to recall the right-hand word. Recall was tested immediately after encoding. Cue words were presented for a maximum duration of five seconds and participants were required to say the pair word aloud. Immediately following an incorrect or a missed trial, the correct response was briefly presented on screen. Participants were instructed to take advantage of this feedback to reinforce their knowledge of the word pair. No feedback was given following a correct trial. If participants recalled fewer than 12 (i.e. <50%) of the word pairs, they then completed an additional recall block. This was repeated a maximum of five times, or until the participant had successfully recalled a minimum of 12 word pairs (whichever was sooner). When participants reached this criterion, they were presented with a final immediate recall block, where they received no feedback on missed or incorrect trials. Finally, delayed recall was tested after a 30-minute delay. Stimuli for this task were taken from the ‘Think/NoThink' procedure used by Anderson and colleagues (Anderson et al., 2004; Benoit & Anderson, 2012) and re-paired to ensure that the paired words had no obvious semantic relation to one another. Due to a software issue only one participant did not complete this.

**Intelligence Quotient Tasks**

***The Wechsler Test of Adult Reading (WTAR; Wechsler, 2001)***

Here participants were shown a list of 50 words and asked to pronounce each word aloud, even if they were unsure of its' correct pronunciation. A single point was awarded for each correctly pronounce word.

### Supplementary Material – Part 3

#### Childhood Emotional Neglect and Wellbeing

**Table S1.** Spearman's correlations and 95% confidence intervals for the relationships between CEN and each of the four ONS wellbeing questions.

| | Cohort 1 | | Cohort 2 | | Combined Cohorts | |
|---|---|---|---|---|---|---|
| | $r_s$ | 95% CI | $r_s$ | 95% CI | $r_s$ | 95% CI |
| Satisfaction | -0.333 | -0.387, -0.275 | -0.310 | -0.386, -0.230 | -0.330 | -0.337, -0.286 |
| Worthwhile | -0.293 | -0.352, -0.235 | -0.329 | -0.409, -0.243 | -0.311 | -0.362, -0.262 |
| Happiness | -0.237 | -0.293, -0.178 | -0.222 | -0.312, -0.133 | -0.236 | -0.287, -0.186 |
| Anxiety | 0.111 | 0.051, 0.171 | 0.060 | -0.038, 0.149 | 0.110 | 0.060, 0.160 |

**Supplementary Material – Part 4**

**ONS Wellbeing Data relative to UK national wellbeing averages**

***Cohort 1: Phase 1***

The mean CEN score for this cohort was 9.12 (SD = 3.84; n = 1027). Overall wellbeing scores ranged from 11 to 40, with a mean of 27.78 (SD = 6.59). We also compared the wellbeing scores of our cohort with the UK national wellbeing averages recorded in the time period July 2014 to June 2015. We therefore compared the mean scores of our cohort, with the mean scores from the participants surveyed in the UK national wellbeing survey, on each of four individual components of the wellbeing scale: i.e. participants' current satisfaction with life [1 (low) - 10 (high)], the extent to which participants believed feel that the things they do in their life are worthwhile [1 (low) - 10 (high)], how happy participants felt yesterday [1 (low) - 10 (high)], and how participants their felt yesterday [1 (low) - 10 (high)]. These comparisons are illustrated in Figure S1. Whilst the average scores of this cohort were lower than the national average on each of the four components, the largest divergence was evident when participants were asked to reflect on their anxiety level on the preceding day. As a group, this undergraduate student population rated their anxiety to be 48.59% higher than that of the UK national average. Life satisfaction, perception that things in their life was worthwhile, and happiness was 2.23%, 9.56% and 7.35% lower than the national average respectively.

***Cohort 2: 2016/2017***

The mean CEN score for this cohort was 9.9 (SD = 4.47; range: 5.0 to 25.0; n = 458). Wellbeing scores ranged from 4.0 to 40.0, with a mean of 24.95 (SD = 6.68). Figure S2 shows Cohort 2 wellbeing scores in relation to the national wellbeing averages between the time period July 2016 and July 2017, which closely reflects the academic year for Cohort 2. Again, it is evident from these average scores, the means of this cohort were markedly lower than that of the UK national average on each of the four components measured. As with Cohort 1, the largest divergence related to anxiety levels. Here this cohort of undergraduate students rated their anxiety yesterday (mean = 5.17 ± 2.52) to be 77.66% higher than that of the UK national average, whilst life satisfaction (mean = 6.62 ± 1.87), perception that things in their life was worthwhile (mean = 6.45 ± 2.13), and happiness yesterday (mean = 6.05 ± 2.23) were 13.91%, 18.04% and 19.55% lower than the national average respectively.

### Supplementary Material – Part 5

**Childhood Emotional Neglect Categories and Depression/Wellbeing**

***Cohort 1: Phase 1***

Participants were categorised by degree of emotional neglect according to the Bernstien and Fink (1997) divisions of "none" (n=631, score 5-10), "low" (n=292, score 11-15), "moderate" (n=70, score 16-20) and "severe" (n=34, score 21-25). A one-way ANOVA revealed a significant difference in overall wellbeing between groups (F=30.257, p<.001). To investigate this finding in more detail, a Tukey HSD bootstrap for multiple comparisons was performed and revealed a significant difference in wellbeing between None-CEN and other groups, and Low-CEN and Severe-CEN. However, no significant difference was observed between Low-CEN and Moderate-CEN, or Moderate-CEN and Severe-CEN (see Table S1). However, when the Life Satisfaction question alone was analysed, a significant difference was observed between all groups.

**Table S2**. Cohort 1 Phase 1 Tukey HSD Bootstrap for Multiple Comparisons

| | Group Comparison | Mean Difference | Confidence Interval |
|---|---|---|---|
| Satisfied | None v Low | 0.731 | $0.505 \leq r_s \leq 0.933$* |
| | None v Moderate | 1.186 | $0.785 \leq r_s \leq 1.609$* |
| | None v Severe | 2.034 | $1.277 \leq r_s \leq 2.792$* |
| | Low v Moderate | 0.455 | $0.032 \leq r_s \leq 0.900$* |
| | Low v Severe | 1.303 | $0.506 \leq r_s \leq 2.081$* |
| | Moderate v Severe | 0.848 | $0.337 \leq r_s \leq 2.081$* |
| Worthwhile | None v Low | 0.817 | $0.549 \leq r_s \leq 1.072$* |
| | None v Moderate | 1.248 | $0.722 \leq r_s \leq 1.820$* |
| | None v Severe | 1.804 | $1.012 \leq r_s \leq 2.716$* |
| | Low v Moderate | 0.432 | $-0.146 \leq r_s \leq 1.020$ |
| | Low v Severe | 0.988 | $0.132 \leq r_s \leq 1.883$* |
| | Moderate v Severe | 0.556 | $-0.423 \leq r_s \leq 1.596$ |
| Happiness | None v Low | 0.791 | $0.466 \leq r_s \leq 1.084$* |
| | None v Moderate | 1.109 | $0.554 \leq r_s \leq 1.677$* |
| | None v Severe | 1.721 | $0.779 \leq r_s \leq 2.677$* |
| | Low v Moderate | 0.318 | $-0.292 \leq r_s \leq 0.893$ |
| | Low v Severe | 0.931 | $-0.010 \leq r_s \leq 1.878$ |
| | Moderate v Severe | 0.613 | $-0.512 \leq r_s \leq 1.671$ |
| Anxiety | None v Low | -0.592 | $-0.965 \leq r_s \leq -0.208$ |
| | None v Moderate | -0.606 | $-1.303 \leq r_s \leq 0.094$ |
| | None v Severe | -0.745 | $-2.013 \leq r_s \leq 0.382$ |
| | Low v Moderate | -0.013 | $-0.726 \leq r_s \leq 0.773$ |
| | Low v Severe | -0.153 | $-1.329 \leq r_s \leq 1.042$ |
| | Moderate v Severe | -0.140 | $-1.494 \leq r_s \leq 1.106$ |
| Overall Wellbeing | None v Low | 2.930 | $2.115 \leq r_s \leq 3.771$* |
| | None v Moderate | 4.241 | $2.710 \leq r_s \leq 6.052$* |
| | None v Severe | 6.478 | $3.933 \leq r_s \leq 9.195$* |
| | Low v Moderate | 1.311 | $-0.434 \leq r_s \leq 3.193$ |
| | Low v Severe | 3.548 | $0.801 \leq r_s \leq 6.306$* |
| | Moderate v Severe | 2.237 | $-1.051 \leq r_s \leq 5.391$ |

*Cohort 2: Phase 1*

When the same statistical analyses were performed on the second cohort based on Bernstein severity categories, a significant difference in wellbeing scores was found (F=10.277, p<.001) with the pattern of differences between groups replicated in the Tukey HSD analysis whereby a significant differences was found between None-CEN and all other groups, between Low-CEN and Severe-CEN but between no other groups (see Figure S4 Table S2).

**Table S3**. Cohort 2 Phase 1. Tukey HSD Bootstrap for Multiple Comparisons.

| | Group Comparison | Mean Difference | Confidence Interval |
|---|---|---|---|
| Satisfied | None v Low | 0.684* | $0.286 \leq r_s \leq 1.110$* |
| | None v Moderate | 1.212* | $0.633 \leq r_s \leq 1.793$* |
| | None v Severe | 1.663* | $0.988 \leq r_s \leq 2.293$* |
| | Low v Moderate | 0.527 | $-0.153 \leq r_s \leq 1.169$ |
| | Low v Severe | 0.978* | $0.267 \leq r_s \leq 1.738$* |
| | Moderate v Severe | 0.451 | $-0.377 \leq r_s \leq 1.276$ |
| Worthwhile | None v Low | 0.820* | $0.368 \leq r_s \leq 1.280$* |
| | None v Moderate | 1.270* | $0.628 \leq r_s \leq 1.899$* |
| | None v Severe | 1.631* | $0.837 \leq r_s \leq 2.357$* |
| | Low v Moderate | 0.451 | $-0.269 \leq r_s \leq 1.175$ |
| | Low v Severe | 0.811* | $0.035 \leq r_s \leq 1.588$* |
| | Moderate v Severe | 0.361 | $-0.592 \leq r_s \leq 1.268$ |
| Happiness | None v Low | 0.739* | $0.260 \leq r_s \leq 1.212$* |
| | None v Moderate | 0.733* | $0.099 \leq r_s \leq 1.370$* |
| | None v Severe | 1.478* | $0.531 \leq r_s \leq 2.387$* |
| | Low v Moderate | -0.007 | $-0.716 \leq r_s \leq 0.767$ |
| | Low v Severe | 0.739 | $-0.245 \leq r_s \leq 1.724$ |
| | Moderate v Severe | 0.746 | $-0.332 \leq r_s \leq 1.769$ |

| | | | |
|---|---|---|---|
| Anxiety | None v Low | -0.449 | $-0.975 \leq r_s \leq 0.065$ |
| | None v Moderate | -0.029 | $-0.735 \leq r_s \leq 0.695$ |
| | None v Severe | -0.394 | $-1.358 \leq r_s \leq 0.698$ |
| | Low v Moderate | 0.421 | $-0.332 \leq r_s \leq 1.226$ |
| | Low v Severe | 0.055 | $-0.952 \leq r_s \leq 1.098$ |
| | Moderate v Severe | -0.365 | $-1.495 \leq r_s \leq 0.704$ |
| Overall Wellbeing | None v Low | 0.673* | $0.347 \leq r_s \leq 1.046$* |
| | None v Moderate | 0.811* | $0.299 \leq r_s \leq 1.294$* |
| | None v Severe | 1.291* | $0.705 \leq r_s \leq 1.870$* |
| | Low v Moderate | 0.137 | $-0.427 \leq r_s \leq 0.706$ |
| | Low v Severe | 0.618 | $-0.001 \leq r_s \leq 1.293$ |
| | Moderate v Severe | 0.481 | $-0.219 \leq r_s \leq 1.257$ |

**Supplementary Material – Part 6**

**Mediation Analysis**

***BDI mediating CEN and EFT SoP***

Direct effect = -0.037 [-0.126, 0.052]

Indirect effect = -0.022 [-0.077, 0.020]

A = 1.243 [0.390, 2.096]

B = -0.018 [-0.057, 0.029]

***BDI mediating CEN and Total EI***

Direct effect = -1.243 [-1.720, -0.765]

Indirect effect = -0.084 [-0.369, .188]

A = 1.243 [0.390, 2.096]

B = -0.068 [-0.273, 0.144]

***BDI mediating CEN and Total Vividness***

Direct effect = -0.044 [-0.099, 0.011]

Indirect effect = -0.016 [-0.051, 0.013]

A = 1.243 [0.390, 2.096]

B = -0.013 [-0.037, 0.012]

***BDI mediating CEN and Total Detail***

Direct effect = -0.044 [-0.099, 0.011]

Indirect effect = -0.016 [-0.051, 0.013]

A = 1.243 [0.390, 2.096]

B = -0.013 [-0.037, 0.012]

***BDI mediating CEN and EFT Vividness***

Direct effect = -0.064 [-0.155, 0.002]

Indirect effect = -0.012 [-0.060, 0.038]

A = 1.243 [0.390, 2.096]

B = -0.010 [-0.053, 0.033]

***BDI mediating CEN and Total Difficulty***

Direct effect = 0.010 [-0.042, 0.062]

Indirect effect = 0.017 [-0.007, 0.053]

A = 1.243 [0.390, 2.096]

B = 0.014 [-0.009, 0.037]
